# Common Substrates of Early Illness Severity: Clinical, Genetic, and Brain Evidence

**DOI:** 10.64898/2026.04.21.26350991

**Authors:** Rochelle Ruby Ye, Clara Vetter, Sidhant Chopra, Stephen Wood, Aswin Ratheesh, Shane Cross, Jente Meijer, Anoja Thanabalasingam, Paris Alexandros Lalousis, Nora Penzel, Linda A. Antonucci, Shalaila S. Haas, Madalina Buciuman, Rachele Sanfelici, Lisa-Maria Neuner, Maria Fernanda Urquijo-Castro, David Popovic, Theresa Lichtenstein, Marlene Rosen, Katharine Chisholm, Alexandra Korda, Georg Romer, Carlo Maj, Anastasia Theodoridou, Anita Riecher-Rössler, Christos Pantelis, Jarmo Hietala, Rebekka Lencer, Alessandro Bertolino, Stefan Borgwardt, Markus Noethen, Paolo Brambilla, Stephan Ruhrmann, Eva Meisenzahl, Raimo K. R. Salokangas, Joseph Kambeitz, Lana Kambeitz-Ilankovic, Peter Falkai, Rachel Upthegrove, Frauke Schultze-Lutter, Nikolaos Koutsouleris, Patrick McGorry, Cassandra Wannan, Barnaby Nelson, Dominic B. Dwyer, PRONIA Consortium

## Abstract

**Background:** The severity of positive psychotic symptoms largely defines emerging psychosis syndromes. However, depressive and negative symptoms are strongly psychologically and biologically interlinked. A transdiagnostic exploration of symptom severity across early illness syndromes could enhance the understanding of shared common factors and future trajectories of mental illness. We aimed to identify subgroups based on the severity of positive, negative, and depressive symptoms and assess relationships with: 1) premorbid functioning, 2) longitudinal illness course, 3) genetic risk, and 4) brain volume differences.

**Methods:** We analysed 749 participants from a multisite, naturalistic, longitudinal (18 months) cohort study of: clinical high risk for psychosis (n=147), recent onset psychosis (n=161), and healthy controls (n=286), and recent onset depression (n=155). Participants were stratified into subgroups based on severity of baseline positive, negative, and depression symptoms. Baseline and longitudinal differences between groups for clinical, functioning, and polygenic risk scores (schizophrenia, depression, cross-disorder) were assessed with ANOVAs and linear mixed models. Voxel-based morphometry was used to examine whole-brain grey matter volume differences. Discovery findings were replicated in a held-out sample (n=610).

**Results:** Participants were stratified into no (n=241), mild (n=50), moderate (n=182), and severe symptom (n=254) subgroups. The mean (SD) age was 25.3 (6.0) and 344 (47.3%) were male. Symptom severity was associated with poorer premorbid functioning and illness trajectory, greater genetic risk, and lower brain volume. Findings were not confounded by the original study groups or symptoms and were largely replicated.

**Conclusions and relevance:** Transdiagnostic symptom severity is linked to shared aetiologies, prognoses, and biological markers across diagnoses and illness stages. Such commonalities could guide therapeutic selection and future research aiming to detect unique contributions to specific psychopathologies.

## Background

Traditional diagnostic systems are limited in guiding research and treatment decisions^1^ because they overlook clinical heterogeneity^2^. Symptom severity is a major contributor, which is measured in established psychosis heuristics by identifying subthreshold (i.e., clinical high risk) and threshold (i.e., first episode psychosis) symptom subgroups^3,4^. When combined with measures of illness course, these are known as stages that are treated with specialised intervention^5–8^. However, heterogeneity extends to severe symptoms in other dimensions, such as depressive and negative symptoms^9^.

Research is progressing towards the use of severity to target care for non-psychotic high-risk^10,11^ and high-acuity^12,13^ cases (e.g., severe depression), using symptom-specific treatments (e.g., antidepressants). However, growing transdiagnostic research suggests that severity across different symptom dimensions^14–17^ may reflect shared etiological factors, such as neurobiological, genetic, or psychosocial mechanisms, resulting in similar clinical trajectories across symptoms and diagnoses^18^. Therefore, transdiagnostic factors such as premorbid functioning, illness course, structural brain changes, or genetic risk may underpin transdiagnostic frameworks^16^ independent of symptom dimension^19^; an argument supported by evidence of increased comorbidity of severe psychopathology^20^.

Epidemiological studies have demonstrated shared genetic risk factors across diagnostic categories^21–23^ with increased risk for a diversity of severe syndromes in the offspring of affected parents^24,25^. Higher schizophrenia polygenic risk scores (PRS), which aggregate the weighted effects of multiple genetic variants associated with schizophrenia, have been associated with increased progression to severe affective and psychotic syndromes, poorer clinical outcomes, cognitive impairment, and increased psychopathology across diagnoses^26–29^. Recent evidence also suggests depression PRS and a new measure of cross-disorder PRS to be a transdiagnostic markers for disorder load^30,31^. Evidence for these two PRS are limited, but recent research has also provided mechanistic support by demonstrating genetic pleiotropy at the cellular level^32^.

Neuroimaging research further supports shared biological substrates across syndromes. Structural brain differences are highly correlated between schizophrenia, depression, and other diagnoses^33,34^ linked to the salience and ventral attention networks^35–37^. Statistical clustering studies corroborate these findings by identifying similar patterns of brain-wide structural differences across syndromes^38^. In similarity to transdiagnostic PRS findings, positive, negative, and depression symptom severity have shown a linear association with lower brain volume^39,40^. However, studies have yet to investigate if transdiagnostic severity is a general factor that mediates these associations.

Other critical gaps also remain in transdiagnostic research. First, there is limited understanding of the clinical antecedents (e.g., premorbid functioning) and longitudinal symptom courses that may differentiate symptom severity groups and inform clinical decisions^41,42^. Second, despite initial evidence that illness courses are largely driven by transdiagnostic symptom severity^16,43,44,^, research has not investigated negative symptom severity, which is suggested to be transdiagnostic^45,46^. Lastly, transdiagnostic genetic and brain differences have not been investigated in early illness groups across symptom severity^47^.

This study aimed to investigate the hypothesis that transdiagnostic symptom severity subgroups are linked to shared premorbid functioning, longitudinal trajectories, PRS, and brain volume patterns. A specific expectation was of increased PRS and decreased brain volume with increasing severity. These hypotheses were investigated and validated in individuals originally classified as clinical high risk for psychosis (CHR-P), recent-onset psychosis (ROP), recent-onset depression (ROD), and a community control group. Secondary analyses were conducted to understand the symptom distribution and correlation across groups, and to ensure that findings were not being driven by diagnosis.

## Methods and Materials

### Participants

1,359 participants were included across 10 sites in Finland, Germany, Italy, Switzerland, and the United Kingdom^48^ (PRONIA). The sample was divided into a discovery set (n=749) recruited between February 2014 and May 2017 (CHR-P, n=147; ROP, n=161; ROD, n=155; HCs, n=286), and an unmatched validation data set (n=610) recruited between May 2017 and July 2019 (CHR-P, n=140; ROP, n=162; ROD, n=130; HCs, n=178)(Table S1). Follow-ups were conducted at 9 and 18 months. Informed consent was obtained from all participants, with assent and parental consent for those younger than 18 years. PRONIA was registered at the German Clinical Trials Register (<u>DRKS00005042</u>) and approved by all local research ethics committees. The PRONIA sample has been used in other publications^38,49,50^.

### Symptom severity groups

Positive and negative symptom severity were assessed using the Structured Interview for Psychosis Risk Syndromes (SIPS; scores 0-6)^51^. Depression symptoms in the last two weeks were measured using the Beck Depression Inventory (BDI-II)^52^. Individuals were stratified into symptom severity groups using SIPS and BDI severity thresholds in correspondence with transdiagnostic clinical staging heuristics (Table 1)^15,51,52^. To accurately represent CHR-P, COGDIS was measured using the Schizophrenia Proneness Instrument (SPI-A)^53^ in addition to positive symptoms. Individuals were reclassified by highest symptom severity (i.e. CHR-P individual with severe depression symptoms are reclassified into the severe group). HCs were also stratified as they may experience mild to moderate symptoms not meeting diagnostic criteria. Individuals with no or questionable symptoms were put in the No Symptoms group.

**Table 1:** Symptom severity score cut-offs for no, mild, moderate, and severe symptom groups.

| Symptom Dimension | No symptoms | Mild symptoms | Moderate symptoms | Severe symptoms |
| --- | --- | --- | --- | --- |
| Depression | BDI-II total score < 14 | BDI-II total score 14-19 | BDI-II score total 20-28 | BDI-II total score > 28 |
| Positive | SIPS positive score < 2 | SIPS positive score = 2<br>OR<br>SIPS positive score = 3, not meeting frequency and duration criteria for moderate symptoms | SIPS positive score = 3-5, occurring at an average frequency of at least once per week for at least several minutes per event in the past month and began within the past year.<sup>1</sup><br>OR<br>SIPS positive score = 6, occurring once in each of the past 3 months for several minutes per day or in the past month for several minutes per day at least once a week (cumulatively more than one hour within the past month), and have recovered spontaneously<sup>2</sup> | SIPS positive score = 6, occurring for at least one day<sup>3</sup> |
| Negative | SIPS negative score < 2 | SIPS negative score = 2 | SIPS negative score 3-5 | SIPS negative score = 6 |
| COGDIS | N/A | N/A | SPI-A score > 2 on at least 2 of 9 cognitive basic symptoms with at least weekly occurrence during the last 3 months<sup>4</sup> | N/A |

Notes: <sup>1</sup>CHR-P UHR criteria for Attenuated Positive Symptoms; <sup>2</sup>CHR-P UHR criteria for Brief Intermittent Psychotic Symptoms; <sup>3</sup>An episode of severe and psychotic positive symptom lasting for at least one day meets DSM-IV-TR diagnostic criteria for a Brief Psychotic Disorder, which is the shortened duration of a psychotic symptom required to be diagnosed with a first episode psychotic disorder<sup>90</sup>. <sup>4</sup>CHR-P COGDIS criteria. BDI-II = Beck Depression Inventory. SIPS = Structured Interview for Psychosis Risk Syndromes

### Clinical symptoms and functioning

Psychological, social, and occupational functioning was measured using the Global Assessment of Functioning scale (GAF)^54^, quality of life measured using the World Health Organisation Quality of Life scale (WHOQoL)^55^, and premorbid functioning at various ages (0-11,12-15, 16-18, >18 years) measured using the Premorbid Adjustment Scale (PAS)^56^.

### Genetic risk

DNA from 619 discovery participants was genotyped using the Illumina Global Screening Array-24 v2 + Psych content^38^. Schizophrenia, depression, and cross-disorder PRS were calculated using the genomic-wide association studies (GWAS) of the Psychiatric Genomics Consortium and the Genomic Psychiatry Cohort^31,57–59^. Computation was completed using PRS-CS, a Bayesian regression framework that infers posterior effect sizes of single nucleotide polymorphisms (SNPs) under a continuous shrinkage prior^60^. We used the PRS-CS auto setting with default hyperparameters to learn the global shrinkage parameter phi from the data using the pre-computed UK Biobank European linkage equilibrium (LD) reference panel supplied with PRS-CS. The first 10 principal components were regressed out to control for population structure.

### Brain imaging

Baseline T1-weighted MRI images were processed using CAT12^61^, including denoising, segmentation, adjusting for inhomogeneity, and registration to the Montreal Neurological Institute template. Grey matter volume (GMV) maps were produced using Jacobian modulation (Methods S2.5). An image quality rating (IQR) score was calculated that combined measures of the noise contrast ratio, inhomogeneity contrast ratio, and image resolution. Participants were excluded if IQR was rated as grade D “Sufficient”. Full imaging procedures are explained in S1.

### Statistical Analyses

#### Clinical analysis

Cases with more than 50% missing values were excluded (n=22; Table S2, S2). Differences in clinical and functioning measures across symptom severity groups were assessed using two-tailed one-way analysis of variance (ANOVA) with false-discovery rate (FDR) corrected post-hoc testing. To ensure robustness^62^, Kruskal-Wallis tests were conducted and presented in the Supplements. Linear mixed effects models were used to examine premorbid functioning and 18-month clinical and functioning trajectories, testing main effects of group and time, linear and quadratic trends, and interactions. Post-hoc tests (estimated marginal means; EMMEANS) were used to compare trends of each measure between groups. Effect sizes (marginal R^2^) were calculated as implemented in the *r2glmm*^63–65^ package in R. Statistical significance was set at FDR-corrected *P* < .05 for multiple comparisons. Full analysis procedure in S2.

#### PRS analysis

Genetic data was not available for 96 individuals, and the mild symptoms group was removed due to small group size (n=43) leaving a sample size of 588. ANOVA with trend analysis was used to identify a linear association in PRS across severity groups. Additional exploratory quadratic analyses were performed for cross-disorder PRS given lack of demonstrated evidence for a linear association compared to schizophrenia and depression PRS. Statistical significance was set at FDR-correct *P* < .05 for multiple comparisons.

#### Brain structure analysis

MRI scans were unavailable for 24 individuals and 4 were excluded due to poor image quality. Additionally, the mild symptoms group was removed due to small group size (n=46) leaving a sample size of 653. Whole brain voxel-based morphometry (VBM) regression was performed using SPM12 running in MATLAB (R2024a) adjusting for age, sex, IQR, and site, with volumes scaled to individuals total intracranial volume (TIV). One-tailed linear contrasts tested the negative association between symptom severity and GMV. Statistical t-maps were enhanced with threshold-free cluster enhancement (TFCE) to improve sensitivity^66^ (https://www.neuro.uni-jena.de/tfce/) and FDR-corrected. Discovery and replication maps were parcellated using a previously validated 100-region cortical^67^ and subcortical atlas^68^ and compared via brain-wide correlations. Statistical significance was assessed using a spatial-autocorrelation preserving permutation test (‘spin-test’)^69^. Anatomical regions were labelled using Automated Anatomical Labelling Atlas 3 (AAL3v1; https://www.oxcns.org/aal3.html)^70^. Full analysis procedure in S1.

#### Replication analyses

The same preprocessing steps to filter participants, and analyses of clinical, genetic, and brain data were conducted in the replication sample and compared with the discovery findings. All measures, variables, and timepoints were the same across samples, with the addition of three new sites (Bari, Dusseldorf, Muenster).

## Results

### Sample characteristics

After filtering, there were 727 participants in the discovery sample (CHR-P, n=143; ROP, n=150; ROD, n=151; HC, n=283). 344 (47.3%) were male and the mean (SD) age was 25.3 (6.0) years. 12 (1.7%) identified as African/Caribbean/Black; 74 (7.4%) identified Asian; 605 (83.2%) identified as European/White; 31 (4.3%) identified as multiracial; and 38 (5.2%) identified with any other racial or ethnic group. After stratification we obtained No (n=241), Mild (n=50), Moderate (n=182), and Severe (n=254) symptom groups. 49 (34%) CHR-P individuals were reclassified into the Severe group. ROD individuals were spread across the Mild (n=21; 14%), Moderate (n=76; 50%), and Severe (n=54; 36%) groups. A limited number of HC individuals were included in the Mild (n=29; 10%) and Moderate (n=12; 4%) subgroups. Full reclassification decisions and baseline clinical differences are presented in Table 2. Kruskal-Wallis tests yielded comparable results with no changes in significant findings (Table S17), and significant associations remain unchanged when controlling for demographic factors (Table S3). Further exploration of study group and symptom distribution, and correlations, are presented in Table S19-22 and S3.

**Table 2:**
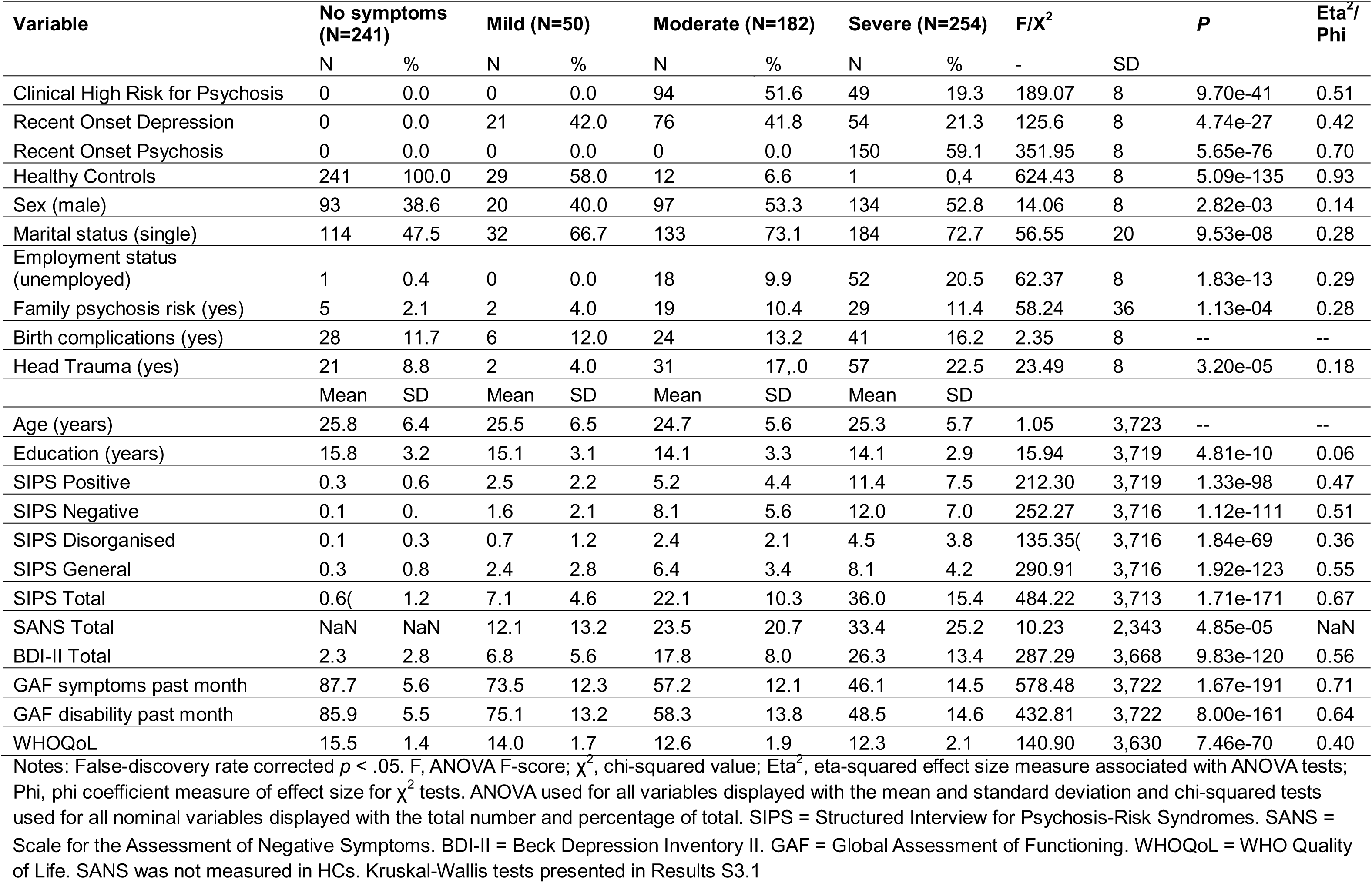
Distribution of original study groups across severity groups and differences across demographic, clinical, and functioning measures.

### Premorbid and current functioning

Individuals with no or mild symptoms showed stable, good premorbid functioning across all developmental stages (ages 0-18+; Figure 1). In contrast, those with moderate or severe symptoms exhibited comparable premorbid functioning in childhood that worsened during youth (*F*_3,2053_ = 16.28, *P* < .001, η = 0.19; Figure 1; Tables S4-5). At baseline, functioning in both symptom (*F*_3,722_ = 578.48, *P* < .001, η^2^ = 0.71) and disability domains (*F*_3,722_ = 432.81, *P* < .001, η^2^ = 0.64) declined with increasing symptom severity.

**Figure 1:**
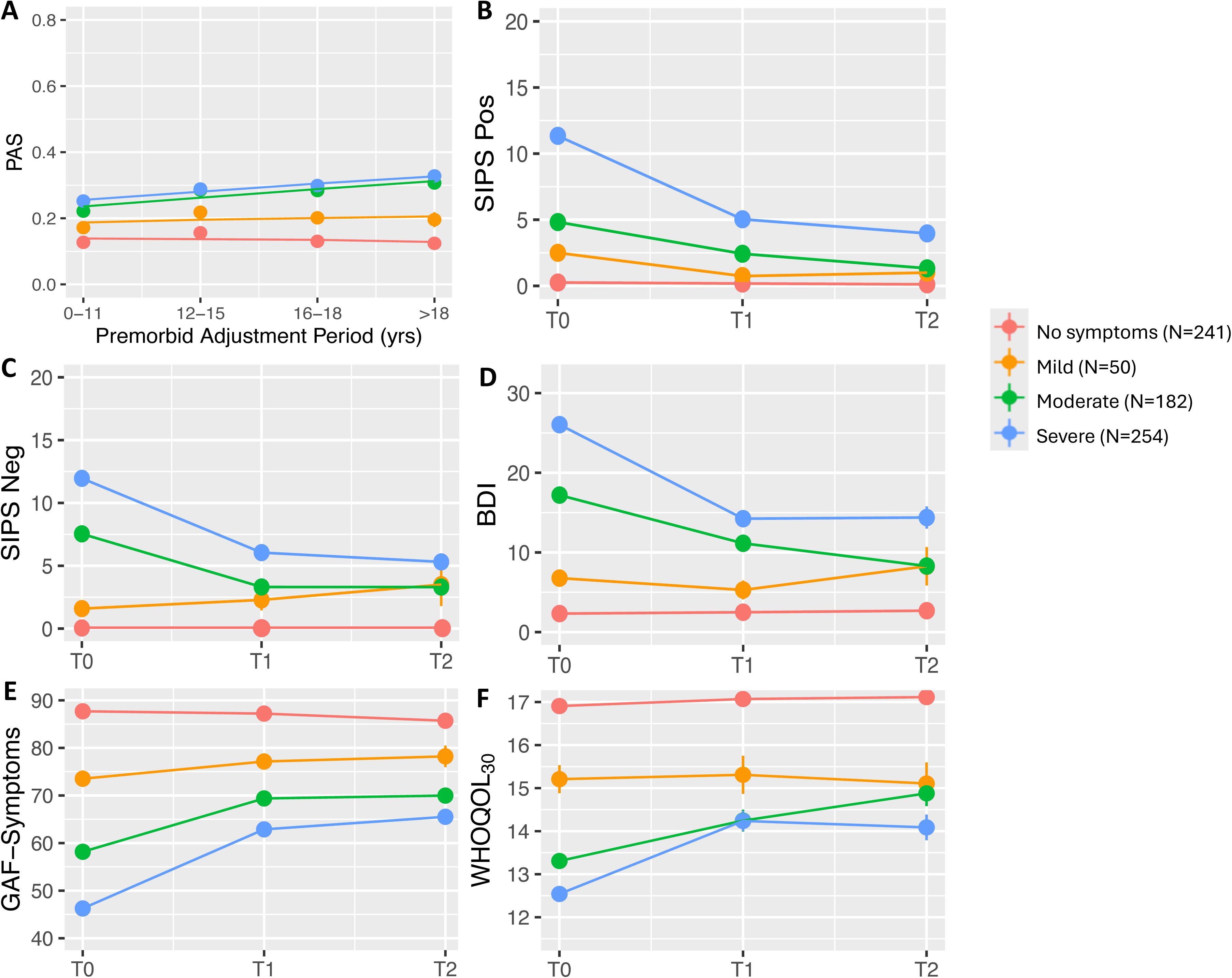
Longitudinal linear-mixed models of A) premorbid adjustment trend from 0 to >18 years for each symptom severity group, and B-F) illness course of groups over 18-months in the discovery sample for B) positive symptoms, C) negative symptoms, D) depression symptoms, E) functioning, and F) quality of life. Notes: Lines represent the fitted predicted values and dots are observed mean (SE) values. T0 = baseline. T1 = 9 months. T2 = 18 months. SIPS = Structured Interview for Psychosis Risk Syndromes. Neg = Negative. Pos = Positive. BDI = Beck Depression Inventory. GAF = Global Assessment of Functioning. WhoQoL_30_ = WHO Quality of Life. Negative symptoms for healthy controls were not assessed during follow up. Alt text: Line graphs showing differences between symptom severity groups on premorbid adjustment, symptoms, global functioning, and quality of life over 18-months.

### Longitudinal Symptoms and Functioning

Of the sample, 64% (466 [174.2]) completed assessments at 9 and 18-months. All participants were included in the longitudinal analyses. Symptom severity show main effects and quadratic group-by-time interactions for SIPS positive (F_3, 861_ = 13.97; *P* < .001, η ^2^ = 0.46), BDI-II (F_3, 755_ = 10.19; *P* < .001, η^2^ = 0.50), GAF disability (F_3, 917_ = 9.79; *P* < .001, η^2^ = 0.56), GAF symptoms (F_3, 915_ = 10.16; *P* < .001, η^2^ = 0.63), and WHOQoL (F_3, 714_ = 5.56; *P* < .001, η^2^ = 0.42; Figure 1). A linear group-by-time interaction was observed for SIPS negative (F_3, 766_ = 9.10; *P* < .001, η ^2^ = 0.42). Full results including post-hoc FDR-corrected trend analyses are presented in Table S6 and Figure S1-2.

To assess whether severity group differences were driven by original study groups, we controlled for ROD and CHR-P (Analysis S1.1). Controlling for ROD and CHR-P, and excluding ROP, had little impact on baseline or longitudinal outcomes (Table S7, Figure S3). Results were also consistent when controlling for symptom dimensions (S4, Figure S15-16, Table S23), suggesting that the findings are robust against diagnosis and symptom.

### PRS

There was a positive linear association between symptom severity and schizophrenia PRS (F_1,584_ = 24.20; P < .001; η² = 0.04) and depression PRS (F_1,584_ = 15.78, P < .001, η² = 0.03). Exploratory analyses revealed a positive quadratic association between symptom severity cross-disorder PRS (F_1,584_ = 8.27, P < .01)(Figure 2) with the moderate symptom group exhibiting the highest PRS. ROP participants played a significant role in the driving the high SCZ-PRS of the severe symptom group (F_2,688_ = 1.56, P = .21, η = 0.0002; Figure S4; S4). Results with the Mild Symptoms group included are presented in Figure S5.

**Figure 2:**
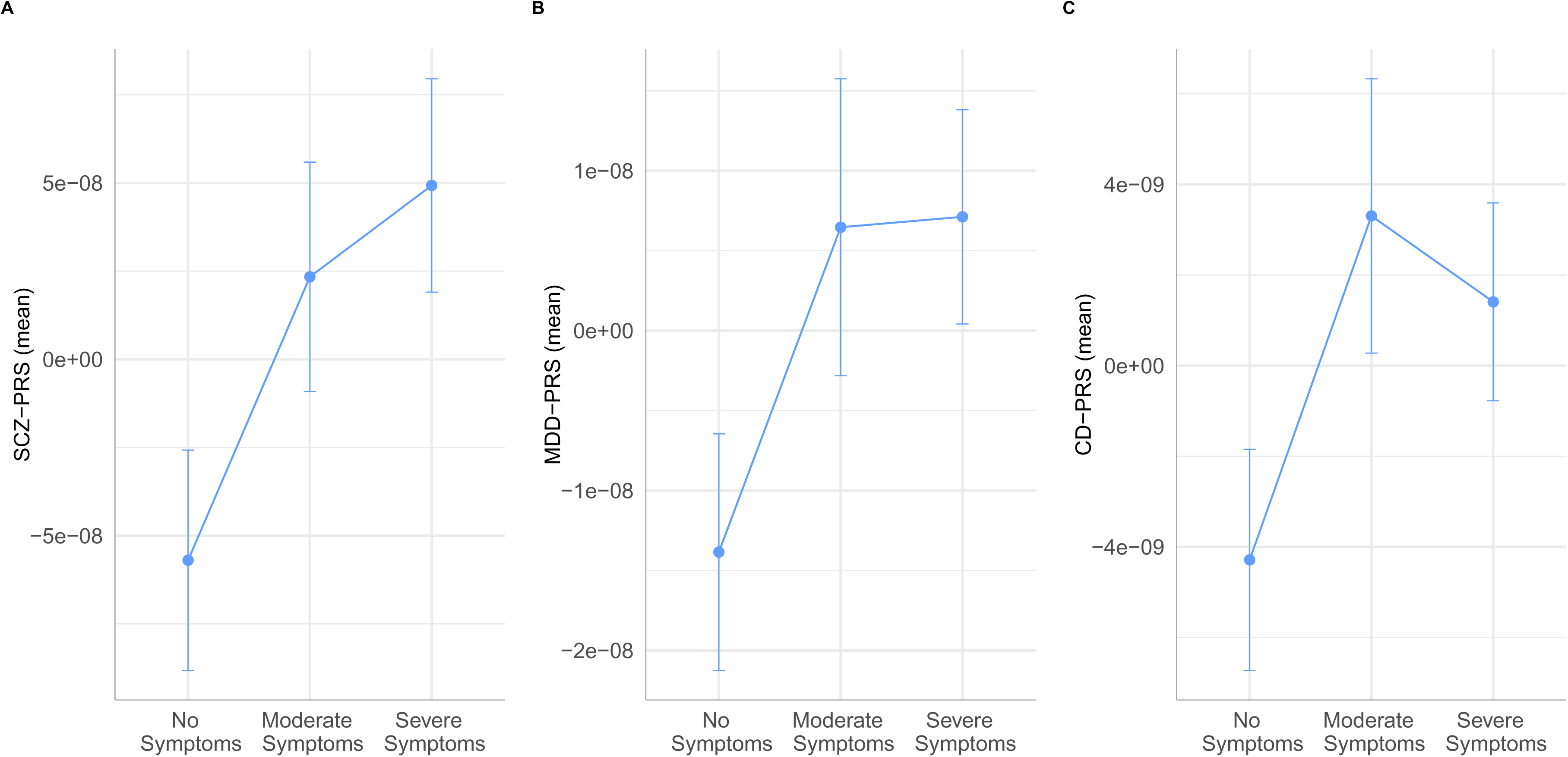
Differences between symptom severity groups on A) schizophrenia, B) depression, and C) cross-disorder PRS Notes: Blue bars show the margin of error about the point estimate. SCZ = Schizophrenia. MDD = Depression. Analysis including the Mild symptom group presented in Figure S5. Alt text: Line graph showing positive linear relationship between symptom severity group, schizophrenia PRS and depression PRS. A quadratic relationship between symptom severity group and cross-disorder PRS. The Mild symptom group is not included

### Brain volume

There was a negative linear relationship between symptom severity and whole-brain GMV (Figure 3), particularly in the bilateral temporal gyrus, middle and anterior cingulate, left insula, and right Rolandic operculum (Table S8. Figure S6-7). This relationship remained after controlling for CHR-P and ROD, with small-to-moderate correlations between discovery and control t-maps (voxel-wise r = .24; parcel-wise r = .14; p < .001; S4; Table S13). Results with the Mild Symptom group included are presented in Figure S8 and Table S10.

**Figure 3:**
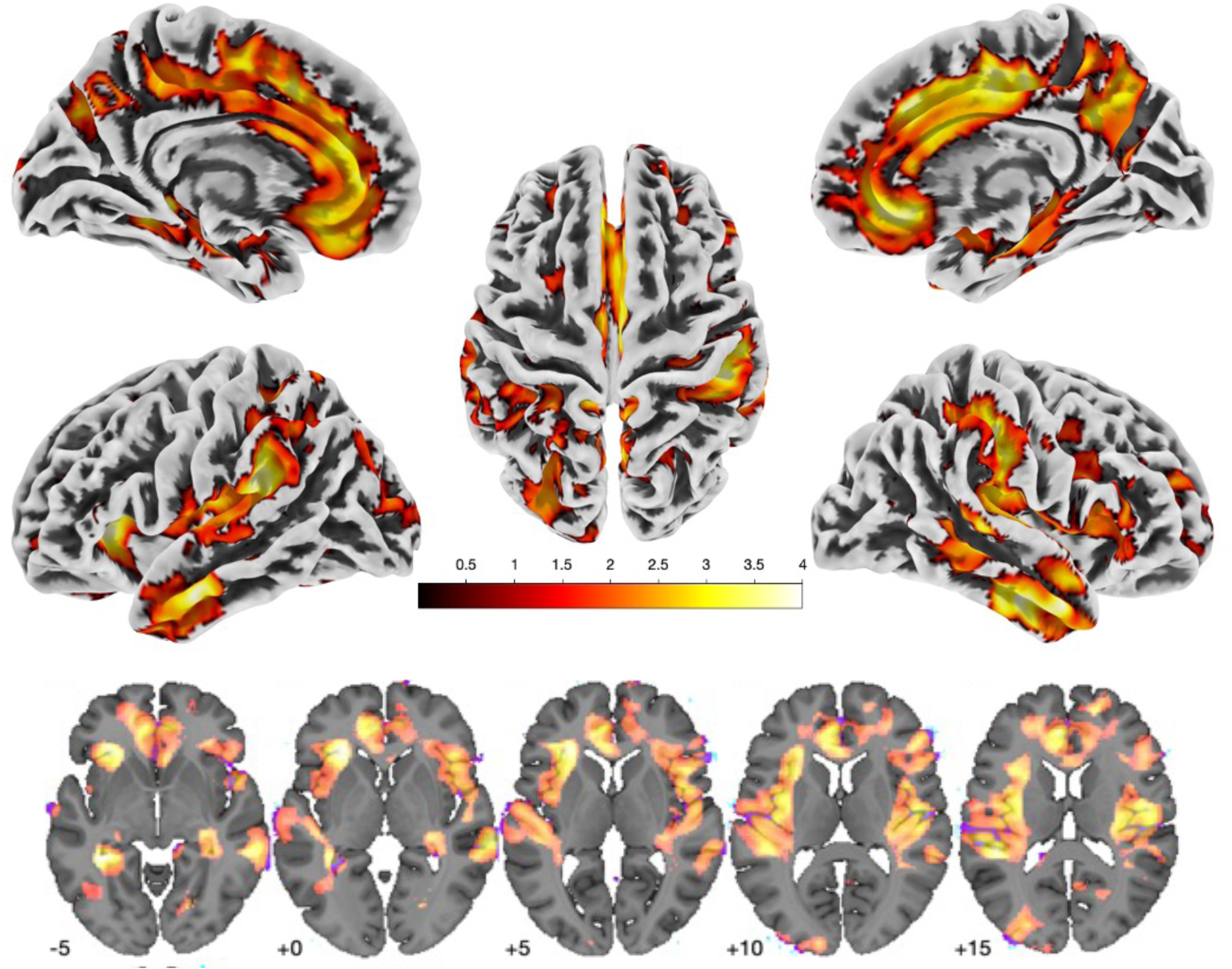
TIV-corrected TFCE surface and subcortical volumes areas of GMV difference across symptom severity groups in the discovery sample. Notes: TIV = Total Intracranial Volume. TFCE = Threshold-Free Cluster Enhancement. FDR-corrected *P <* .05. Analysis including the Mild group presented in Figure S8. Alt text: Brain scans depicting statistical map of GMV differences identified via voxel-based morphometry and threshold-free cluster enhancement, overlaid with a canonical map

### Replication

Of the 610 participants in the replication sample, 45 were removed due having more than 50% missing data, leaving a final sample of 565 (CHR-P, n = 132; ROP, n = 145; ROD, n = 124; HC, n = 164). The mean (SD) age was 24.8 (5.7) and 265 (47%) were male. 488 (86.4%) identified as European/White, 19 (3.36) identified as Asian, 15 (2.65%) identified as African/Caribbean/Black, 15 (2.65%) identified as multiracial, and 25 (4.42%) identified as any other racial or ethnic group.

Clinical and functioning findings were replicated at baseline and longitudinally (Table S11-12, Figure S9-10). Kruskal-Wallis tests yielded comparable results (Table S18). Symptom severity groups also exhibited similar premorbid functioning trends as the discovery sample (*F*_3,1516_ = 8.51, *P* < .001, η^2^ = 0.21; Figure S11, Table S13-14). Group-level genetic analyses were replicated for schizophrenia and depression PRS (Figure S12, Table S15). Cross-disorder PRS showed similar association patterns, however, the ANOVA analysis did not reach significance. VBM statistical maps from the discovery and replication samples were moderately correlated (voxel *r* = .21, parcel *r* = .44, *p* < .05; Table S16, Figure S13-14).

## Discussion

Our findings extend previous transdiagnostic research by demonstrating that symptom severity in early-stage psychopathology is reflected across clinical, genetic, and neuroanatomical domains simultaneously. While prior studies have largely examined single modalities (e.g., brain volume) or focussed on established illness^25,35,38,71^, the present study investigated whether a transdiagnostic severity dimension spanning CHR-P, ROP, ROD, and HCs corresponds to shared biological differences in the early stages of emerging illness. Study groups were distributed across stages, highlighting that symptom burden cuts across diagnostic boundaries. Importantly, we found that premorbid functioning, PRS, and GMV differed markedly in parallel with transdiagnostic symptom severity. These findings suggest that symptom severity captures a shared liability dimension that cuts across traditional diagnostic categories, rather than reflecting disorder-specific illness progression.

Greater symptom severity was associated with poorer premorbid functioning, particularly during early adolescence, highlighting the importance of early intervention irrespective of diagnosis^72^. Longitudinally, while all groups showed symptom improvement over time, individuals with greater baseline symptom severity remained significantly impaired clinically and functionally over 18 months^38^. These findings indicate that transdiagnostic symptom severity captures clinically meaningful differences in illness trajectory, supporting dimensional approaches that move beyond syndrome-specific frameworks. This aligns with emerging models of transdiagnostic staging that conceptualise mental disorders along shared severity dimensions and may help guide clinical decision making across diagnostic categories^73–75^.

Aligned with the hypotheses, schizophrenia PRS was positively associated with symptom severity, explaining around 4% of variance. While previous research has demonstrated elevated schizophrenia PRS across diagnostic groups^26,29,76^, the present findings extend this work by showing that polygenic liability marks transdiagnostic symptom severity during the early stages of emerging illness, independent of diagnostic labels^77,78^. This suggests that genetic liability related to schizophrenia may indicate broader vulnerability processes that manifest across syndromes rather than mapping exclusively onto psychotic disorders^79^. Depression and cross-disorder PRS were also positively associated with symptom severity, consistent with literature demonstrating substantial polygenic overlap between psychiatric disorders^31^. Interestingly, we observed a quadratic relationship between cross-disorder PRS and symptom severity, with the highest scores observed in the moderate symptom group. This pattern may reflect the sample characteristics used to calculate the cross-disorder PRS^30^. To our knowledge, this represents that first investigation of the relationship between cross-disorder PRS and transdiagnostic symptom severity, prompting further investigation to clarify these findings.

Brain volume analyses further supported the hypothesis of transdiagnostic mechanisms. We observed a linear pattern of lower GMV across groups in the middle temporal gyrus, cingulate cortex, and insula, consistent with known transdiagnostic brain signatures^34,35^. Importantly, these findings were independent of study group, suggesting that the observed structural differences reflect neurobiological correlates of symptom severity rather than disorder-specific pathology^38,49,80,81^. The areas identified are key components of the salience network and default node network, which are involved in processing reward, motivation, emotion, and social cognition^82^, functions often disrupted across both psychotic and affective disorders. Notably, the neuroanatomical patterns were detectable in early illness stages, reinforcing their potential as early intervention targets. In addition, the observed GMV-based pattern may provide a useful framework for controlling transdiagnostic severity differences in future studies seeking to isolate symptom-specific mechanisms (e.g., positive symptoms).

Taken together, the convergence of clinical course, genetic liability, and neurobiological differences across symptom severity suggests that transdiagnostic symptom severity captures a biologically meaningful dimension of illness. While it may be expected that greater symptom severity is associate with poorer functioning, our findings demonstrate that this clinical association is also paralleled by genetic and neurobiological differences across diagnoses. This supports the idea that symptom severity reflects a shared liability dimension that can be detected in the early stages of emerging illness, rather being simply a clinical descriptor.

### Limitations

Several limitations should be considered. First, the sample was recruited into single-syndrome groups (i.e. depression, psychosis), which may confound transdiagnostic analyses, although we controlled for diagnosis in supplementary analysis to ensure robustness of our findings. Future transdiagnostic studies could be designed to further investigate the findings herein with a range of diagnoses and illness stages; e.g., because positive, negative, and depressive symptoms are prevalent in conditions such as bipolar^83,84^ and borderline personality disorder^85,86^.

Secondly, we used the total score for positive symptoms to define severity, however, an earlier study on CHR-P symptoms indicated that suspiciousness/persecutory ideas (SIPS positive item P2) be regarded as a transdiagnostic feature uniquely^19^. Future studies should examine transdiagnostic and disorder-specific severity markers on a symptom level to elucidate important features.

Thirdly, COGDIS was underrepresented in this sample. These symptoms may follow a non-linear course and be underreported during acute phases^87^. COGDIS symptoms have been reported to be largely psychosis-specific^19^ and related to a schizophrenia-specific brain signature^88^.

Finally, our study was limited to three symptom dimensions and CHR-P recruitment may have reduced the representation of milder positive symptoms. Broader recruitment and inclusion of additional cardinal symptom dimensions (e.g. from the Hierarchical Taxonomy of Psychopathology)^89^ would enhance the study of severity across syndromes.

## Conclusion

Our findings provide multimodal evidence that transdiagnostic symptom severity in early-stage illness corresponds to shared genetic liability and neurobiological differences, independent of diagnostic-label. These findings support the notion that psychiatric disorders may share underlying mechanisms that influence illness course and presentation across disorders. By demonstrating linear associations between symptom severity, polygenic risk, and GMV differences, the study highlights potential limitations of disorder-specific staging frameworks that may obscure broader dimensions of illness burden. Future research should further investigate transdiagnostic severity models to clarify how shared and disorder-specific processes contribute to the development and progression of psychiatric disorders.

## Disclosures

Dr Koutsouleris, Dr Ruhrmann, Dr Riecher-Rossler report grants from European Union during the conduct of the study. Dr Koutsouleris and Dr Meisenzahl hold issued patent US20160192889A1 (‘Adaptive pattern recognition for psychosis risk modelling’). Dr Sanfelici reports personal fees from Lundbeck Australia Pty Ltd outside the submitted work. Dr Hietala reports personal fees from Orion ltd, personal fees from Otsuka, personal fees from Lundbeck, and other from Takeda during the conduct of the study. Dr Rössler reports grants from The Zurich Program for Sustainable Development of Mental Health Services (ZInEP) was supported by a private donation. The donor had no further role in the experimental design, collection, analysis, interpretation of data, writing, and submitting this paper for publication during the conduct of the study. Dr Pantelis reports grants from Australian NHMRC during the conduct of the study; personal fees from Lundbeck Australia Pty Ltd outside the submitted work. Dr Upthegrove reports personal fees from Sunovion outside the submitted work. No other disclosures were reported.

## Data Availability

All data produced in the present study are available upon reasonable request to the authors

## Acknowledgments

PRONIA is a Collaboration Project funded by the European Union under the 7th Framework Programme under grant agreement n° 602152. David Popovic was supported by the Else-Kröner-Fresenius-Foundation through the Clinician Scientist Program ‘EKFS-Translational Psychiatry’. Theresa Lichtenstein supported through the Koeln Fortune Program/ Faculty of Medicine, University of Cologne (No 370/2020). Dominic Dwyer was supported by a NARSAD Young Investigator grant no. 30196.

## Role of the Funders

The funders were not involved in the design and conduct of the study; collection, management, analysis, and interpretation of the data; preparation, review, or approval of the manuscript; and decision to submit the manuscript for publication.

## The PRONIA consortium

The authors listed here performed the screening, recruitment, rating, examination, and follow-up of the study participants. They were involved in implementing the examination protocols of the study, setting up its IT infrastructure, and organizing the flow and quality control of the data analyzed in this manuscript between the local study sites and the central study database.

*Department of Psychiatry and Psychotherapy, Ludwig-Maximilian-University, Munich, Germany*

Shalaila Haas, Alkomiet Hasan, Claudius Hoff, Ifrah Khanyaree, Aylin Melo, Susanna Muckenhuber-Sternbauer, Yanis Köhler, Ömer Öztürk, Nora Penzel, David Popovic, Adrian Rangnick, Sebastian von Saldern, Rachele Sanfelici, Moritz Spangemacher, Ana Tupac, Maria Fernanda Urquijo, Johanna Weiske, Antonia Wosgien, Camilla Krämer

*Department of Psychiatry and Psychotherapy, University of Cologne, Cologne, Germany*

Karsten Blume, Dennis Hedderich, Dominika Julkowski, Nathalie Kaiser, Thorsten Lichtenstein, Ruth Milz, Alexandra Nikolaides, Tanja Pilgram, Mauro Seves, Martina Wassen

*Department of Psychiatry (Psychiatric University Hospital, UPK), University of Basel, Switzerland*

Christina Andreou, Laura Egloff, Fabienne Harrisberger, Ulrike Heitz, Claudia Lenz, Letizia Leanza, Amatya Mackintosh, Renata Smieskova, Erich Studerus, Anna Walter, Sonja Widmayer

*Institute of Mental Health & School of Psychology, University of Birmingham, United Kingdom*

Chris Day, Sian Lowri Griffiths, Mariam Iqbal, Mirabel Pelton, Pavan Mallikarjun, Alexandra Stainton, Ashleigh Lin

*Department of Psychiatry, University of Turku, Finland*

Alexander Denissoff, Anu Ellilä, Tiina From, Markus Heinimaa, Tuula Ilonen, Päivi Jalo, Heikki Laurikainen, Antti Luutonen, Akseli Mäkela, Janina Paju, Henri Pesonen, Reetta-Liina Säilä, Anna Toivonen, Otto Turtonen

*Department of Psychiatry (Psychiatric University Hospital LVR/HHU Düsseldorf), University of Düsseldorf, Germany*

Sonja Botterweck, Norman Kluthausen, Gerald Antoch, Julian Caspers, Hans-Jörg Wittsack

*Department of Basic Medical Science, Neuroscience and Sense Organs - University of Bari Aldo Moro*

Giuseppe Blasi, Giulio Pergola, Grazia Caforio, Leonardo Fazio, Tiziana Quarto, Barbara Gelao, Raffaella Romano, Ileana Andriola, Andrea Falsetti, Marina Barone, Roberta Passiatore, Marina Sangiuliano.

*Department of Psychiatry and Psychotherapy of the University of Münster, Germany*

Marian Surmann, Olga Bienek, Udo Dannlowski

*General Electric Global Research Inc., USA*, Ana Beatriz Solana, Manuela Abraham, Timo Schirmer

*Workgroup of Paolo Brambilla, University of Milan, Italy*

- Department of Neuroscience and Mental Health, Fondazione IRCCS Ca’ Granda Ospedale Maggiore Policlinico, University of Milan, Milan, Italy: Carlo Altamura, Marika Belleri, Francesca Bottinelli, Adele Ferro, Marta Re
- Programma2000, Niguarda Hospital, Milan: Emiliano Monzani, Maurizio Sberna
- San Paolo Hospital, Milan: Armando D’Agostino, Lorenzo Del Fabro
- Villa San Benedetto Menni, Albese con Cassano (CO): Giampaolo Perna, Maria Nobile, Alessandra Alciati

*Workgroup of Paolo Brambilla at the University of Udine, Italy*

- Department of Medical Area, University of Udine, Udine, Italy: Matteo Balestrieri, Carolina Bonivento, Giuseppe Cabras, Franco Fabbro
- IRCCS Scientific Institute “E. Medea”, Polo FVG, Udine: Marco Garzitto, Sara Piccin

## Access to data and data analysis

Rochelle Ruby Ye had full access to all the data in the study, performed the entire data analysis (except for the brain and genetic data preprocessing) and thus takes the responsibility for the integrity of the data and the accuracy of the data analysis. Franziska Degenhardt, Carlo Maj, Oleg Borisov and Peter Krawitz were responsible for genotyping and computation of the polygenic risk scores and thus take responsibility for this part of the analysis.

## Notes

### Author Declarations

PRONIA was registered at the German Clinical Trials Register (DRKS00005042). Ethikkommission der Medical Fakultat der LMU gave ethics approval for all sites

