## Supplementary Material for "Common Substrates of Early Illness Severity: Clinical, Genetic, and Brain Evidence"

[Figure S6: Discovery *t*-statistical maps showing areas of GMV differences across symptom severity groups using a linear contrast (*p* < .001, uncorr.).	21](#_Toc227335251)

[Figure S8: Discovery *t*-statistical maps showing areas of GMV differences across symptom severity groups including the Mild group using a linear contrast (*p* < .001, uncorr.).	23](#_Toc227335253)

[Figure S13: Replication *t*-statistical maps showing areas of GMV differences across symptom severity groups using a linear contrast (*p* < .001, uncorr.)	29](#_Toc227335258)

### Table S1: Study inclusion / exclusion criteria of the study. Reproduced from Dwyer, et al. ^1^

| **Group Inclusion Criteria** | | **Group Exclusion Criteria** | **General Inclusion / Exclusion Criteria** |
| --- | --- | --- | --- |
| **Clinical High Risk (CHR)** | | | **Inclusion Criteria:**  1. Age 15 to 40 years  2. Language skills sufficient for participation  3. Able to provide to consent / assent  **Exclusion Criteria:**  1. IQ below 70  2. Hearing is not sufficient for neurocognitive testing  3. Current or past head trauma with loss of consciousness (> 5 min)  4. Current or past known neurological disorder of the brain  5. Current or past known somatic disorder potentially affecting the structure or functioning of the brain  6. Current or past alcohol dependence  7. Current polysubstance dependence or within the past six months (Note: any combination with E.6. led to exclusion)  8. Any contra-indication for MRI  **Exclusion criteria for healthy controls:**  1. Any current or past DSM-IV axis disorder  2. A positive familial history (1st degree relatives) for affective or non-affective psychoses or major affective disorders; and  3. An intake of psychotropic medications or drugs more than 5 times/year and in the month before study inclusion.  **Drop-out criteria:**  1. No follow-up examination after the 6-months follow-up examination (IV6)  2. Withdrawn consent / assent |
| **Psychosis-risk syndrome defined:**  **EITHER** by *Attenuated Positive Symptoms (APS),* as measured by the SIPS (requires 1 of 5 attenuated psychotic symptoms: unusual thought content/ delusional ideas, suspiciousness/persecutory ideas, grandiosity, perceptual abnormalities/hallucinations, and disorganized communication) with a moderate to severe, but not psychotic, severity (SIPS score 3-5) that (1) began with-in the past year or was rated one or more scale points higher compared to 12 month ago, **AND** (2) occurred at an average frequency of at least once per week for at least several minutes per event in the past month | | Any intake of antipsychotic medication for more than 30 cumulative days at or above the minimum dosage threshold defined by the DGPPN S3 Guidelines for the treatment of first-episode psychosis ^2^.  Any intake of antipsychotic drugs within the past 3 months before psychopathological baseline assessments at or above the minimum dosage threshold.  Occurrence of the CHR syndrome is better explained by other DSM-IV disorder |  |
| **OR:** by *Brief Intermittent Psychotic Symptoms (BLIPS),* as measured by the SIPS (as defined by one of the symptoms listed above (1) reaching a psychotic level of intensity in each of the past 3 months for at least several minutes per day, **OR** (2) reaching a psychotic level of intensity in the past month, occurring at an average frequency of at least once per week for at least several minutes per event in the past month, or occurring at least for a cumulative period of more than one hour within the past month, **AND** (1+2) remitting spontaneously within one week (i.e. without antipsychotic medication) | |  |  |
| **OR:** by a *Genetic Risk and Functional Decline Psychosis-Risk Syndrome (GRFD)* defined by a current 30% or greater reduction in the functional disability score of the split version of the Global Assessment of Functioning Scale (GAF-F) compared with the highest lifetime level of functioning, **AND** (having a first-degree relative with a history of any psychotic disorder, **OR** having a DSM-IV-TR schizotypal personality disorder). | |  |  |
| **OR:** by a Cognitive Disturbance Syndrome (COGDIS) as measured by the SPI-A (requires at least 2 of 9 cognitive basic symptoms with at least weekly occurrence (score ≥3) during the last 3 months) | |  |  |
| **Recent Onset Depression (ROP)** | | |  |
| **Recent-onset Depression as defined by DSMIV-TR major depression category + all of the following criteria:**  1. First life-time depressive episode,  2. Duration of current depressive episode no longer than 24 months,  3. Diagnostic criteria fulfilled within past three months | 1. Occurrence of the major depressive episode is better explained by other DSM-IV disorder  2. See CHR exclusion criteria | |  |
| **Recent Onset Psychosis (ROP)** | | |  |
| **Psychotic episode as defined by DSM-IV-TR affective or non-affective psychotic episode category + all of the following criteria:**  1. Psychotic episode in the past 3 months.  2. Onset of psychosis within the past 24 months | Antipsychotic medication intake longer than 90 cumulative days with a daily dose rate at or above the minimum dosage threshold defined as by the DGPPN S3 Guidelines for the treatment of first-episode psychosis^2^. | |  |

### Table S2: Comparisons of the filtered discovery sample with excluded cases

| **Variable** | **Discovery** | **50% Missing** | **P-value** |
| --- | --- | --- | --- |
| N | 727 | 22 |  |
| Clinical High Risk, N(%) | 143(19.7) | 4(18.2) | -- |
| Recent Onset Depression, N(%) | 151(20.8) | 4(18.2) | -- |
| Recent Onset Psychosis, N(%) | 150(20.6) | 11(50.0) | 9.55e-04 |
| Control Participants, N(%) | 283(38.9) | 3(13.6) | -- |
| Age, yrs(SD) | 25.3(6.0) | 24.5(6.3) | -- |
| Gender, male(%) | 344(47.3) | 14(63.6) | -- |
| Education, yrs(SD)) | 14.7(3.2) | 9.7(2.9) | -- |
| Marital status, single(%) | 463(64.0) | 2(50.0) | -- |
| Employment status, unemployed(%) | 71(9.8) | 1(4.5) | -- |
| Family psychosis risk, yes(%) | 55(7.6) | 1(9.1) | -- |
| Birth complications, yes(%) | 99(13.7) | 2(14.3) | -- |
| Head Trauma <5min LOC, yes(%) | 111(15.3) | 4(28.6) | -- |
| SIPS Positive, mean(SD) | 5.5(6.8) | 7.8(5.8) | -- |
| SIPS Negative, mean(SD) | 6.3(7.2) | 0(0.0) | -- |
| SIPS Disorganised, mean(SD) | 2.2(3.1) | 0(0.0) | -- |
| SIPS General, mean(SD) | 4.7(4.6) | 0(0.0) | -- |
| SIPS Total, mean(SD) | 18.8(18.3) | -- | -- |
| SANS Total, mean(SD) | 23.9(22.1) | 30.0(0.0) | -- |
| BDI Total, mean(SD) | 14.8(13.8) | 7.8(9.4) | -- |
| GAF symptoms past month, mean(SD) | 64.5(21.1) | 58.1(17.8) | -- |
| GAF disability past month, mean(SD) | 65.1(20.0) | 62.5(16.3) | -- |
| WHO QoL Total | 13.6(2.3) | 14.5(0.9) | -- |
| WAIS-V Vocabulary | 11.1(3.1) | 9.6(2.9) | -- |
| WAIS-V Matrix Reasoning | 10.7(2.6) | 9.2(1.0) | -- |

Notes: False discovery rate corrected *p-*values show

**Table S3: Baseline demographic, clinical, and functioning measures across severity groups controlling for demographics**

| **Variable** | **No symptoms (N=239)** | **Mild (N=48)** | **Moderate (N=181)** | **Severe (N=253)** | **F/X^2^** | ***P*** | **Eta^2^/Phi** |
| --- | --- | --- | --- | --- | --- | --- | --- |
|  | **N** |  |  |  |  |  |  |
| Sex (male) | 92 | 20 | 97 | 134 | 5.83 | 1.31e-01 | 0.09 |
| Marital status (single) | 114 | 32 | 132 | 184 | 34.24 | 7.06e-04 | 0.11 |
| Employment status (unemployed) | 1 | 0 | 18 | 52 | 48.44 | 2.74e-10 | 0.26 |
| Family psychosis risk (yes) | 5 | 2 | 18 | 29 | 62.09 | 4.06e-05 | 0.1 |
| Birth complications (yes) | 28 | 6 | 24 | 41 | 3.09 | 3.79e-01 | 0.07 |
| Head trauma (yes) | 21 | 2 | 31 | 57 | 20.81 | 1.39e-04 | 0.17 |
|  | **Mean** |  |  |  |  |  |  |
| Age (years) | 20.62 | 21.76 | 21.49 | 21.68 | 1.62 | 1.92e-01 | 0.01 |
| Education (years) | 13.6 | 12.84 | 12.34 | 12.35 | 24.15 | 1.17e-14 | 0.09 |
| SIPS Positive | 1.46 | 3.47 | 5.69 | 11.73 | 219.14 | 6.13e-100 | 0.48 |
| SIPS Negative | 2.61 | 3.87 | 9.61 | 12.98 | 283.91 | 3.38e-120 | 0.55 |
| SIPS Disorganised | 0.98 | 1.59 | 3.04 | 5.04 | 138.66 | 2.86e-70 | 0.37 |
| SIPS General | 0.65 | 2.8 | 6.69 | 8.38 | 288.27 | 2.14e-121 | 0.55 |
| SIPS Total | 5.38 | 11.47 | 24.74 | 37.89 | 531.46 | 4.61e-179 | 0.69 |
| SANS Total | NA | 22.39 | 26.9 | 35.4 | 17.44 | 8.81e-08 | 0.09 |
| BDI Total | 0.88 | 5.7 | 17.25 | 25.99 | 288.54 | 8.53e-119 | 0.57 |
| GAF Social past month | 82.69 | 69.68 | 54.02 | 43.65 | 604.89 | 1.94e-193 | 0.72 |
| GAF Disability past month | 80.11 | 70.81 | 55.05 | 46.17 | 474.6 | 4.77e-168 | 0.67 |
| WHOQoL Total | 15.47 | 14.16 | 12.74 | 12.37 | 146 | 4.14e-71 | 0.42 |
| WAIS-V Vocabulary | 10.96 | 10.64 | 10.55 | 10.26 | 11.9 | 1.86e-07 | 0.05 |
| WAIS-V Matrix Reasoning | 10.94 | 11.21 | 10.92 | 10.33 | 8.65 | 1.66e-05 | 0.04 |

Notes: Analysis of covariance (ANCOVA) with sex, age, education, marital status, and employment status as covariates. False-discovery rate corrected p < .05. F, ANOVA F-score; χ2, chi-squared value; Eta2, eta-squared effect size measure for ANOVA tests; Phi, phi coefficient measure of effect size for χ2 tests. SIPS = Structured Interview for Psychosis-Risk Syndromes. SANS = Scale for the Assessment of Negative Symptoms. BDI-II = Beck Depression Inventory II. GAF = Global Assessment of Functioning. WHOQoL = WHO Quality of Life. SANS was not measured in HCs.

### Table S4: PAS linear mixed model analysis

| **term** | **meansq** | **df** | **DenDF** | **F** | **p** | **sig** | **Rsq** |
| --- | --- | --- | --- | --- | --- | --- | --- |
| D | 0.9374411 | 3 | 712.1297 | 84.33423 | < 0.001 | *** |  |
| time | 0.4389872 | 1 | 2,054.5744 | 39.49224 | < 0.001 | *** |  |
| D:time | 0.1809149 | 3 | 2,053.3017 | 16.27550 | < 0.001 | *** |  |
| R-squared |  |  |  |  |  |  | 0.19 |

Notes: D = severity group; meansq = mean square error; DenDF = denominator degrees of freedom; F = F statistic; Rsq = R-squared. Degrees of freedom approximated using Satterthwaite method.

### Table S5: Pairwise estimated marginal means in linear PAS course

| **contrast** | **estimate** | **SE** | **df** | **t.ratio** | **p.value** | **signif** |
| --- | --- | --- | --- | --- | --- | --- |
| D1 - D2 | -0.6237381 | 0.4707467 | 2,055.792 | -1.3249973 | 0.22 |  |
| D1 - D3 | -1.7149220 | 0.2933605 | 2,055.154 | -5.8457829 | < 0.001 | *** |
| D1 - D4 | -1.6084483 | 0.2684190 | 2,049.721 | -5.9923049 | < 0.001 | *** |
| D2 - D3 | -1.0911839 | 0.4843573 | 2,057.910 | -2.2528493 | 0.049 | * |
| D2 - D4 | -0.9847103 | 0.4696704 | 2,056.311 | -2.0965984 | 0.054 | . |
| D3 - D4 | 0.1064737 | 0.2916304 | 2,056.491 | 0.3650981 | 0.72 |  |

Notes: D1 = no symptoms; D2 = mild symptoms; D3 = moderate symptoms; D4 = severe symptoms; SE = standard mean error; df = degrees of freedom; t.ratio = t-score ratio

### Table S6: Mixed model analysis of symptom and functioning course over 18-months

| **term** | **meansq** | **NumDF** | **DenDF** | **F** | **p** | **sig** | **Rsq** |
| --- | --- | --- | --- | --- | --- | --- | --- |
| **SIPS Positive** |  |  |  |  |  |  | 0.46 |
| D | 1,450.6027 | 3 | 637.5410 | 138.09642 | < 0.001 | *** |  |
| time | 1,192.2539 | 1 | 934.3410 | 113.50178 | < 0.001 | *** |  |
| time^2 | 251.7384 | 1 | 868.5397 | 23.96533 | < 0.001 | *** |  |
| D:time | 698.3179 | 3 | 931.6401 | 66.47941 | < 0.001 | *** |  |
| D:time^2 | 146.6057 | 3 | 861.0935 | 13.95676 | < 0.001 | *** |  |
| **SIPS Negative** |  |  |  |  |  |  | 0.42 |
| D | 3,937.35865 | 3 | 887.0394 | 240.325448 | < 0.001 | *** |  |
| time | 348.98139 | 1 | 805.8468 | 21.300856 | < 0.001 | *** |  |
| time^2 | 139.97280 | 1 | 704.8676 | 8.543551 | 0.004 | ** |  |
| D:time | 149.17683 | 2 | 766.2483 | 9.105340 | < 0.001 | *** |  |
| D:time^2 | 19.01086 | 2 | 675.7368 | 1.160370 | 0.31 |  |  |
| **BDI** |  |  |  |  |  |  | 0.50 |
| D | 6,969.9278 | 3 | 656.7084 | 163.46763 | < 0.001 | *** |  |
| time | 2,222.3646 | 1 | 822.5278 | 52.12173 | < 0.001 | *** |  |
| time^2 | 777.5870 | 1 | 755.8059 | 18.23696 | < 0.001 | *** |  |
| D:time | 2,014.1326 | 3 | 821.1631 | 47.23801 | < 0.001 | *** |  |
| D:time^2 | 434.5339 | 3 | 755.8875 | 10.19124 | < 0.001 | *** |  |
| **GAF-DI** |  |  |  |  |  |  | 0.56 |
| D | 18,292.2607 | 3 | 720.1656 | 248.287532 | < 0.001 | *** |  |
| time | 8,241.0965 | 1 | 977.2409 | 111.859411 | < 0.001 | *** |  |
| time^2 | 1,841.4694 | 1 | 920.6269 | 24.994936 | < 0.001 | *** |  |
| D:time | 5,459.2514 | 3 | 975.1689 | 74.100412 | < 0.001 | *** |  |
| D:time^2 | 721.3257 | 3 | 917.5159 | 9.790817 | < 0.001 | *** |  |
| **GAF-S** |  |  |  |  |  |  | 0.63 |
| D | 28,376.9834 | 3 | 697.2445 | 374.97780 | < 0.001 | *** |  |
| time | 8,738.7802 | 1 | 985.4947 | 115.47558 | < 0.001 | *** |  |
| time^2 | 2,289.3013 | 1 | 919.2916 | 30.25118 | < 0.001 | *** |  |
| D:time | 5,894.3676 | 3 | 982.6839 | 77.88907 | < 0.001 | *** |  |
| D:time^2 | 769.2742 | 3 | 915.5215 | 10.16531 | < 0.001 | *** |  |
| **WHOQoL** |  |  |  |  |  |  | 0.42 |
| D | 221.597771 | 3 | 682.8485 | 129.872947 | < 0.001 | *** |  |
| time | 43.230799 | 1 | 757.3037 | 25.336497 | < 0.001 | *** |  |
| time^2 | 23.047397 | 1 | 711.9884 | 13.507507 | < 0.001 | *** |  |
| D:time | 31.505601 | 3 | 758.2252 | 18.464650 | < 0.001 | *** |  |
| D:time^2 | 9.492611 | 3 | 713.6869 | 5.563383 | < 0.001 | *** |  |

Notes: D = severity group; meansq = mean square error; NumDF = numerator degrees of freedom; DenDF = denominator degrees of freedom; F = F statistic; Rsq = R-squared

### Table S7: Comparison of severity groups across demographic, clinical, and functioning measures. Discovery and replication samples combined (n = 997) and ROP diagnostic group removed.

| **Variable** | **No symptoms** | **Mild** | **Moderate** | **Severe** | **F/Chi2** | **Sig** | **Eta2/Phi** |
| --- | --- | --- | --- | --- | --- | --- | --- |
| N | 392 | 74 | 308 | 223 |  |  |  |
| Clinical High Risk, N(%) | 0(0.0) | 0(0.0) | 159(51.6) | 116(52.0) | 333.27(8) | 6.25e-72 | 0.58 |
| Recent Onset Depression, N(%) | 0(0.0) | 36(48.6) | 134(43.5) | 105(47.1) | 247.31(8) | 2.50e-53 | 0.50 |
| Recent Onset Psychosis, N(%) | 0(0.0) | 0(0.0) | 0(0.0) | 0(0.0) | NaN(4) | -- | -- |
| Control Participants, N(%) | 392(100.0) | 38(51.4) | 15(4.9) | 2(0.9) | 856.55(8) | 2.35e-185 | 0.93 |
| Age, yrs(SD) | 25.4(5.9) | 25.3(6.2) | 24.7(5.7) | 24.3(5.9) | 2.19(3,993) | -- | -- |
| Gender, male(%) | 157(40.1) | 28(37.8) | 162(52.6) | 93(41.7) | 13.76(12) | 3.24e-02 | 0.12 |
| Education, yrs(SD)) | 15.8(3.1) | 15.0(3.0) | 14.0(3.0) | 13.8(2.7) | 32.30(3,984) | 6.43e-20 | 0.09 |
| Marital status, single(%) | 182(46.5) | 49(69.0) | 218(70.8) | 149(67.4) | 63.86(20) | 4.43e-09 | 0.25 |
| Employment status, unemployed(%) | 240(63.8) | 34(49.3) | 148(49.5) | 102(47.9) | 62.65(12) | 1.30e-11 | 0.25 |
| Family psychosis risk, yes(%) | 8(2.1) | 3(4.1) | 42(13.7) | 31(14.1) | 93.77(44) | 1.74e-08 | 0.31 |
| Birth complications, yes(%) | 42(10.7) | 11(14.9) | 51(16.6) | 45(20.5) | 11.28(8) | 1.03e-02 | 0.11 |
| Head Trauma, yes(%) | 31(7.9) | 6(8.1) | 49(15.9) | 42(19.1) | 20.11(8) | 1.61e-04 | 0.14 |
| SIPS Positive, mean(SD) | 0.2(0.6) | 2.2(2.1) | 5.2(4.7) | 5.5(4.8) | 160.83(3,984) | 8.02e-85 | 0.33 |
| SIPS Negative, mean(SD) | 0.0(0.2) | 1.9(2.3) | 8.7(5.5) | 12.1(6.4) | 442.87(3,982) | 6.52e-182 | 0.58 |
| SIPS Disorganised, mean(SD) | 0.0(0.3) | 0.8(1.3) | 2.5(2.2) | 3.2(3.0) | 164.16(3,979) | 3.40e-86 | 0.33 |
| SIPS General, mean(SD) | 0.2(0.7) | 2.7(3.1) | 6.9(3.6) | 9.1(3.8) | 565.33(3,981) | 2.83e-213 | 0.63 |
| SIPS Total, mean(SD) | 0.5(1.1) | 7.6(5.5) | 23.2(10.7) | 29.9(13.1) | 676.18(3,971) | 3.14e-237 | 0.68 |
| SANS Total, mean(SD) | NaN(NaN) | 10.9(11.3) | 20.5(18.6) | 28.7(23.4) | 15.33(2,471) | 3.56e-07 | NaN |
| BDI Total, mean(SD) | 2.4(2.9) | 7.5(5.5) | 18.5(7.8) | 33.6(10.4) | 959.15(3,911) | 2.47e-281 | 0.76 |
| GAF social past month, mean(SD) | 87.8(5.6) | 72.4(12.7) | 55.0(12.7) | 51.0(12.6) | 821.63(3,991) | 3.36e-268 | 0.71 |
| GAF disability past month, mean(SD) | 86.5(5.6) | 74.2(13.5) | 55.7(14.1) | 50.8(14.1) | 628.84(3,991) | 8.31e-229 | 0.66 |
| WHO QoL Total | 15.5(1.4) | 13.8(2.0) | 12.6(1.8) | 11.5(1.8) | 297.37(3,876) | 4.29e-133 | 0.50 |
| WAIS-V Vocabulary | 12.2(2.8) | 11.2(2.9) | 10.9(2.9) | 11.2(4.1) | 10.43(3,969) | 9.30e-07 | 0.03 |
| WAIS-V Matrix Reasoning | 11.5(2.2) | 11.1(2.5) | 10.7(2.6) | 11.1(4.0) | 4.59(3,926) | 3.37e-03 | 0.01 |

Notes: F, ANOVA F-score; χ2 , chi-squared value; df, degrees of freedom; p-value, p-value associated with ANOVA or χ2 (only false-discovery rate values shown); Eta2 , eta-squared effect size measure associated with ANOVA tests; Phi, phi coefficient measure of effect size for χ2 tests. ANOVA used for all variables displayed with the mean and standard deviation [mean(SD)] and chi-squared tests used for all nominal variables displayed with the

### Table S8: Discovery VBM findings in the three-group linear trend analysis.

|  | **Anatomical region** | **k (number of voxels)** |
| --- | --- | --- |
| 56; -9; -26 | Right middle temporal gyrus | 1171 |
| 9; -9; 38 | Right middle cingulate | 994 |
| -60; -8; -22 | Left middle temporal gyrus | 487 |
| 8; 33; -14 | Right superior frontal gyrus | 345 |
| -28; 27; 0 | Left insula | 499 |
| 52; -26; 42 | Right supramarginal gyrus | 344 |
| -51; -36; 22 | Left superior temporal gyrus | 632 |
| -3; 12; 42 | Left middle cingulate | 108 |
| 50; -10; 12 | Right Rolandic operculum | 141 |
| -8; 39; 16 | Left anterior cingulate cortex-pregenual | 69 |

Notes: VBM = Voxel-based morphometry; Only clusters with k > 50 are listed. Anatomical regions labelled using Automated anatomical labelling atlas 3 (AAL3)^3^. Uncorrected p <.001.

### Table S9: VMB findings in the three-group linear trend analysis controlling for CHR and ROD, removing ROP

| **Coordinates for maximum voxel** | **Anatomical region** | **k (number of voxels)** |
| --- | --- | --- |
| 14; -27; 34 | Right middle cingulate | 77 |
| 26; -64; 0 | Right lingual gyrus | 82 |
| 18; -14; -16 | Right hippocampus | 79 |
| -33; -34; 45 | Left postcentral gyrus | 105 |
| -15; -70; -44 | Left cerebellum | 75 |

Notes: VBM = Voxel-based morphometry; Only clusters with *k* > 50 are listed. Anatomical regions labelled using Automated anatomical labelling atlas 3 (AAL3)^3^. Uncorrected *p* <.001.

### Table S10: VMB findings in the four-group linear trend analysis

| **Coordinates for maximum voxel** | **Anatomical region** | **k (number of voxels)** |
| --- | --- | --- |
| 21; -20; 21 | Right parahippocampus | 2592 |
| 56; -10; -27 | Right middle temporal gyrus | 648 |
| 46; -16; 40 | Right postcentral gyrus | 271 |
| 9; 32; -14 | Right superior frontal gyrus-medial orbita | 142 |
| -26; -86; 15 | Left middle occipital gyrus | 94 |
| -27; 22; -3 | Left Insula | 406 |
| 6; 24; 34 | Right middle cingulate & paracingulate gyri | 59 |
| 50; -10; 14 | Right Rolandic operculum | 75 |

Notes: VBM = Voxel-based morphometry; Only clusters with *k* > 50 are listed. Anatomical regions labelled using Automated anatomical labelling atlas 3 (AAL3)^3^. Uncorrected *p* <.001.

### Table S11: Comparison of symptom severity groups in the replication sample (n = 565) across demographic, clinical, and functioning measures

| **Variable** | **No symptoms** | **Mild** | **Moderate** | **Severe** | **F/Chi2** | **Sig** | **Eta2/Phi** |
| --- | --- | --- | --- | --- | --- | --- | --- |
| N | 151 | 24 | 126 | 264 |  |  |  |
| Clinical High Risk, N(%) | 0(0.0) | 0(0.0) | 65(51.6) | 67(25.4) | 110.01(8) | 1.09e-23 | 0.44 |
| Recent Onset Depression, N(%) | 0(0.0) | 15(62.5) | 58(46.0) | 51(19.3) | 109.23(8) | 1.61e-23 | 0.44 |
| Recent Onset Psychosis, N(%) | 0(0.0) | 0(0.0) | 0(0.0) | 145(54.9) | 222.40(8) | 6.09e-48 | 0.63 |
| Control Participants, N(%) | 151(100.0) | 9(37.5) | 3(2.4) | 1(0.4) | 518.64(8) | 4.35e-112 | 0.96 |
| Age, yrs(SD) | 25.0(5.0) | 24.7(5.7) | 24.8(5.9) | 24.7(6.0) | 0.05(3,561) | -- | -- |
| Gender, male(%) | 64(42.4) | 8(33.3) | 65(51.6) | 128(48.5) | 4.99(12) | -- | -- |
| Education, yrs(SD)) | 15.9(3.1) | 14.9(3.0) | 13.8(2.7) | 14.1(7.5) | 4.30(3,555) | 5.16e-03 | 0.02 |
| Marital status, single(%) | 68(45.0) | 17(73.9) | 85(67.5) | 198(75.6) | 49.54(16) | 1.31e-07 | 0.30 |
| Employment status, unemployed(%) | 99(71.2) | 10(43.5) | 56(45.9) | 111(44.6) | 43.85(12) | 7.91e-08 | 0.28 |
| Family psychosis risk, yes(%) | 3(2.0) | 1(4.2) | 23(18.4) | 39(14.9) | 75.31(36) | 3.33e-07 | 0.37 |
| Birth complications, yes(%) | 14(9.3) | 5(20.8) | 27(21.4) | 57(21.9) | 11.41(8) | 9.69e-03 | 0.14 |
| Head Trauma, yes(%) | 10(6.6) | 4(16.7) | 18(14.3) | 53(20.3) | 14.08(8) | 2.80e-03 | 0.16 |
| SIPS Positive, mean(SD) | 0.1(0.4) | 1.5(1.7) | 5.3(5.2) | 11.9(7.4) | 151.29(3,552) | 1.50e-71 | 0.45 |
| SIPS Negative, mean(SD) | 0.0(0.2) | 2.5(2.7) | 9.5(5.3) | 11.8(7.0) | 162.48(3,551) | 1.93e-75 | 0.47 |
| SIPS Disorganised, mean(SD) | 0.0(0.2) | 1.2(1.4) | 2.6(2.3) | 4.1(3.8) | 69.79(3,544) | 3.40e-38 | 0.28 |
| SIPS General, mean(SD) | 0.1(0.5) | 3.2(3.8) | 7.6(3.9) | 8.0(4.5) | 162.90(3,545) | 2.27e-75 | 0.47 |
| SIPS Total, mean(SD) | 0.3(0.9) | 8.5(7.2) | 24.8(11.1) | 35.9(14.9) | 310.61(3,535) | 9.95e-117 | 0.64 |
| SANS Total, mean(SD) | NaN(NaN) | 12.1(13.2) | 23.5(20.7) | 33.4(25.2) | 10.23(2,343) | 4.85e-05 | NaN |
| BDI Total, mean(SD) | 2.6(3.2) | 8.9(5.4) | 19.5(7.5) | 25.8(13.5) | 163.63(3,491) | 1.62e-73 | 0.50 |
| GAF symptoms past month, mean(SD) | 87.9(5.6) | 70.0(13.4) | 51.7(12.8) | 44.4(14.4) | 426.28(3,560) | 4.15e-144 | 0.70 |
| GAF disability past month, mean(SD) | 87.6(5.7) | 72.3(14.2) | 51.8(13.7) | 46.3(13.9) | 392.99(3,560) | 2.53e-137 | 0.68 |
| WHO QoL Total | 15.4(1.5) | 13.4(2.4) | 12.6(1.6) | 12.1(2.1) | 100.27(3,469) | 3.72e-50 | 0.39 |
| WAIS-V Vocabulary | 12.6(2.6) | 10.8(2.7) | 10.9(3.0) | 10.3(4.1) | 14.33(3,537) | 5.36e-09 | 0.07 |
| WAIS-V Matrix Reasoning | 12.0(2.0) | 10.8(2.3) | 10.6(2.7) | 10.5(4.0) | 7.87(3,538) | 3.81e-05 | 0.04 |

Notes: F, ANOVA F-score; χ2 , chi-squared value; df, degrees of freedom; p-value, p-value associated with ANOVA or χ2 (only false-discovery rate values shown); Eta2 , eta-squared effect size measure associated with ANOVA tests; Phi, phi coefficient measure of effect size for χ2 tests. ANOVA used for all variables displayed with the mean and standard deviation [mean(SD)] and chi-squared tests used for all nominal variables displayed with the total number and percentage of total [n(%)].

### Table S12: Comparison of filtered discovery and replication sample across clinical, cognitive, brain, and medication use data.

| **Variable** | **Discovery** | **Replication** | **F/Chi2** | **Sig** | **Eta2/Phi** |
| --- | --- | --- | --- | --- | --- |
| N | 727 | 565 |  |  |  |
| Clinical High Risk, N(%) | 143(19.7) | 132(23.4) | 2.59(4) | -- | -- |
| Recent Onset Depression, N(%) | 151(20.8) | 124(21.9) | 0.26(4) | -- | -- |
| Recent Onset Psychosis, N(%) | 150(20.6) | 145(25.7) | 4.57(4) | -- | -- |
| Control Participants, N(%) | 283(38.9) | 164(29.0) | 13.77(4) | 2.06e-04 | 0.10 |
| Age, yrs(SD) | 25.3(6.0) | 24.8(5.7) | 1.55(1290) | -- | -- |
| Gender, male(%) | 344(47.3) | 265(46.9) | 6.46(6) | -- | -- |
| Education, yrs(SD)) | 14.7(3.2) | 14.5(5.6) | 0.79(1280) | -- | -- |
| Marital status, single(%) | 463(64.0) | 368(65.5) | 5.98(10) | -- | -- |
| Employment status, unemployed(%) | 71(9.8) | 73(12.9) | 3.19(4) | -- | -- |
| Family psychosis risk, yes(%) | 55(7.6) | 66(11.8) | 16.64(22) | -- | -- |
| Birth complications, yes(%) | 99(13.7) | 103(18.4) | 5.29(4) | 2.15e-02 | 0.06 |
| Head Trauma <5min LOC, yes(%) | 111(15.3) | 85(15.1) | 0.01(4) | -- | -- |
| SIPS Positive, mean(SD) | 5.5(6.8) | 6.8(7.6) | -3.09(1277) | 2.05e-03 | 0.01 |
| SIPS Negative, mean(SD) | 6.3(7.2) | 7.7(7.4) | -3.32(1273) | 9.36e-04 | 0.01 |
| SIPS Disorganised, mean(SD) | 2.2(3.1) | 2.5(3.3) | -1.75(1266) | -- | -- |
| SIPS General, mean(SD) | 4.7(4.6) | 5.6(5.0) | -3.28(1267) | 1.07e-03 | 0.01 |
| SIPS Total, mean(SD) | 18.8(18.3) | 22.3(18.9) | -3.32(1254) | 9.32e-04 | 0.01 |
| SANS Total, mean(SD) | 23.9(22.1) | 29.6(24.2) | -3.27(721) | 1.11e-03 | 0.01 |
| BDI Total, mean(SD) | 14.8(13.8) | 17.2(14.2) | -2.85(1165) | 4.39e-03 | 0.01 |
| GAF symptoms past month, mean(SD) | 64.5(21.1) | 58.8(22.1) | 4.78(1288) | 1.96e-06 | 0.02 |
| GAF disability past month, mean(SD) | 65.1(20.0) | 59.7(21.5) | 4.70(1288) | 2.88e-06 | 0.02 |
| WHO QoL Total | 13.6(2.3) | 13.2(2.3) | 2.62(1105) | 8.87e-03 | 0.01 |
| WAIS-V Vocabulary | 11.1(3.1) | 11.1(3.6) | -0.02(1243) | -- | -- |
| WAIS-V Matrix Reasoning | 10.7(2.6) | 10.9(3.3) | -1.50(1196) | -- | -- |

Notes: All results are FDR-corrected *p* <.05. Differences in the replication appear driven by a lower recruitment of healthy controls, resulting in overall higher scores in clinical symptomatology. mean(SD).

### Table S13: PAS linear mixed model analysis in the replication sample

| **term** | **meansq** | **df** | **DenDF** | **F** | **p** | **sig** | **Rsq** |
| --- | --- | --- | --- | --- | --- | --- | --- |
| D | 0.8590993 | 3 | 528.3457 | 70.966555 | < 0.001 | *** |  |
| time | 0.2015758 | 1 | 1,514.1458 | 16.651319 | < 0.001 | *** |  |
| D:time | 0.1030679 | 3 | 1,516.2871 | 8.515002 | < 0.001 | *** |  |
| R-squared |  |  |  |  |  |  | 0.21 |

Notes: D = severity group; meansq = mean square error; DenDF = denominator degrees of freedom; F = F statistic; Rsq = R-squared; degrees of freedom approximated using Satterthwaite method.

### Table S14: Pairwise estimated marginal means in linear PAS course in the replication sample

| **contrast** | **estimate** | **SE** | **df** | **t.ratio** | **p.value** | **signif** |
| --- | --- | --- | --- | --- | --- | --- |
| D1 - D2 | 0.4583311 | 0.6090375 | 1,515.370 | 0.7525499 | 0.45 |  |
| D1 - D3 | -0.5113930 | 0.3350609 | 1,518.995 | 1.5262689 | .15 |  |
| D1 - D4 | -1.2918912 | 0.2857283 | 1,524.350 | 4.5213977 | < 0.001 | *** |
| D2 - D3 | -0.9697241 | 0.6192569 | 1,515.421 | 1.5659479 | 0.15 |  |
| D2 - D4 | -1.7502223 | 0.5940152 | 1,516.350 | 2.9464269 | 0.01 | ** |
| D3 - D4 | 0.7804984 | 0.3069090 | 1,523.361 | 2.5430929 | 0.02 | * |

Notes: D1 = no symptoms; D2 = mild symptoms; D3 = moderate symptoms; D4 = severe symptoms; SE = standard mean error; df = degrees of freedom; t.ratio = t-score ratio; Rsq = R-squared

### Table S15: Comparison of symptom severity groups in the replication sample across PRS scores.

| **Variable** | **No** | **Moderate** | **Severe** | **F/Chi2** | **Sig** | **Eta2/** |
| --- | --- | --- | --- | --- | --- | --- |
| N | 141 | 111 | 243 |  |  |  |
| SCZ-PRS | -5.45e-08  (-1.4e-07) | -1.00e-08  (2.06e-07) | 3.83e-08  (2.22e-07) | 8.79(2,490) | 1.77e-04 | 0.03 |
| MDD-PRS | -1.32e-08  (5.32e-08) | 5.76e-10  (4.86e-08) | 6.27e-09  (5.26e-08) | 6.30(2,490) | 1.99e-03 | 0.03 |
| Cross-Disorder PRS | -1.69e-09  (1.73e-08) | 6.76e-10  (1.76e-08) | 3.48e-10  (1.67e-08) | 0.81(2,490) | -- | -- |

SCZ = Schizophrenia, MDD = Depression. Mean(SD)

### Table S16: Replication VBM findings in the three-group linear trend analysis.

| **Coordinates for maximum voxel** | **Anatomical region** | **k (number of voxels)** |
| --- | --- | --- |
| 51; -24; 15 | Right superior temporal gyrus | 1145 |
| 45; 15; -3 | Right insula | 543 |
| 2; 45; -24 | Right gyrus rectus | 362 |
| -63;-36; 21 | Left superior temporal gyrus | 130 |

Notes: VBM = Voxel-based morphometry; Only clusters with *k* > 50 are listed. Anatomical regions labelled using Automated anatomical labelling atlas 3 (AAL3)^3^. Uncorrected *p* <.001.

### Figure S1: Symptom severity group comparison on clinical measures of depressive, positive, and negative symptoms, self-reported functioning, and clinician-reported quality of life in the discovery sample.

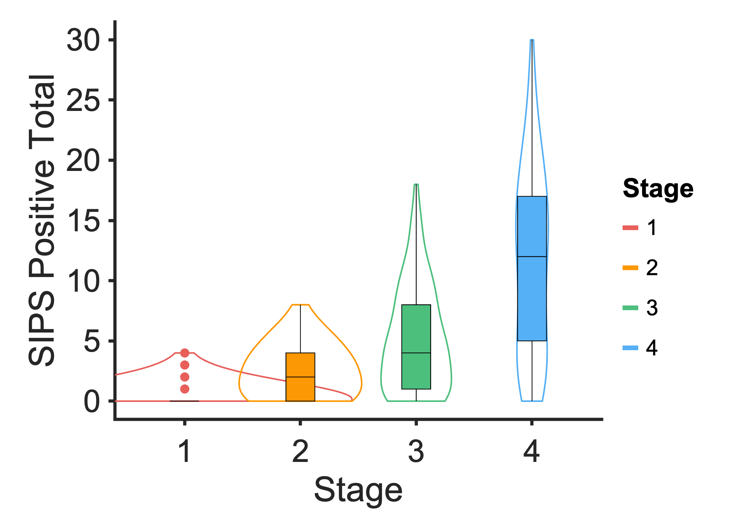

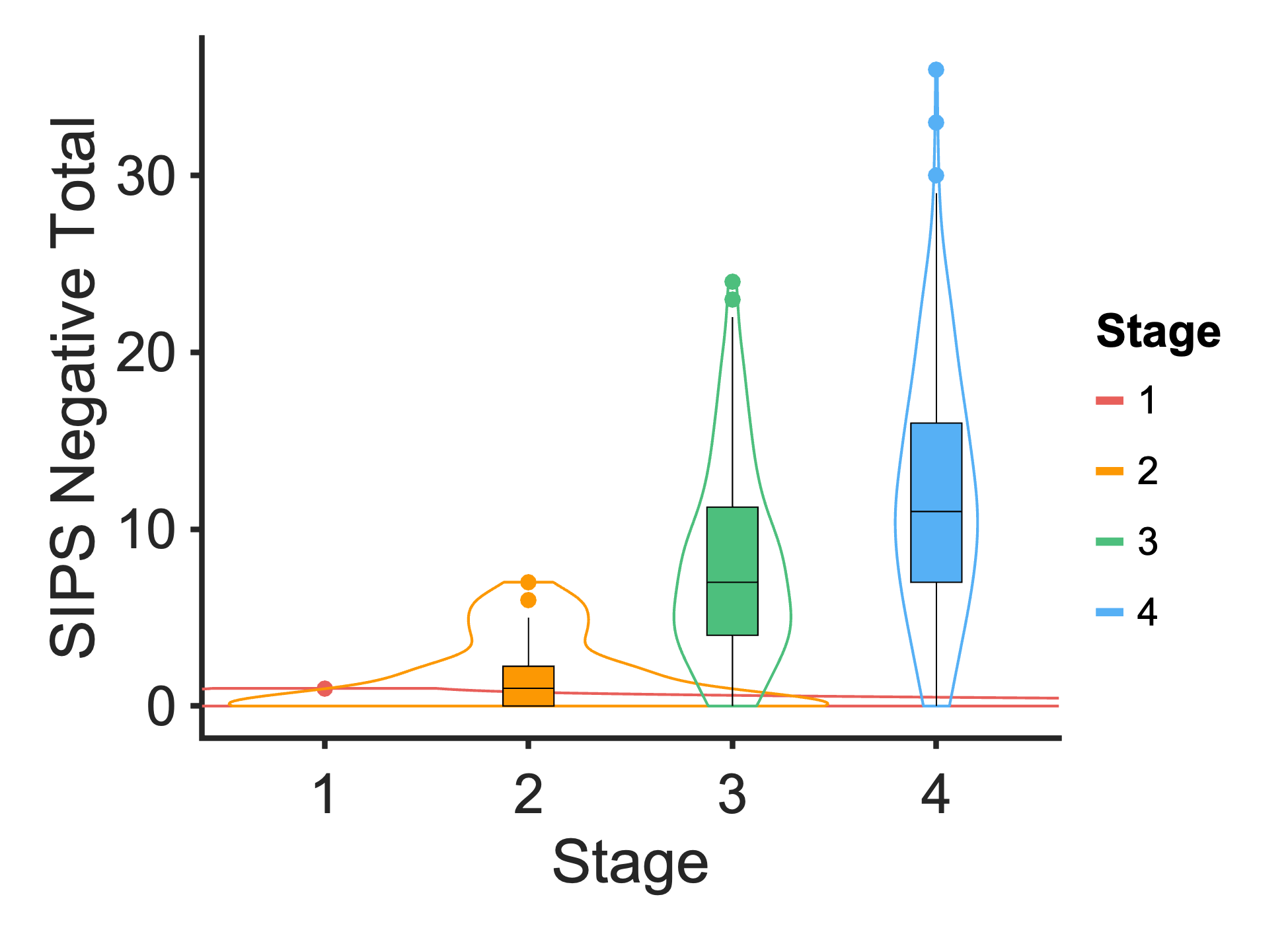

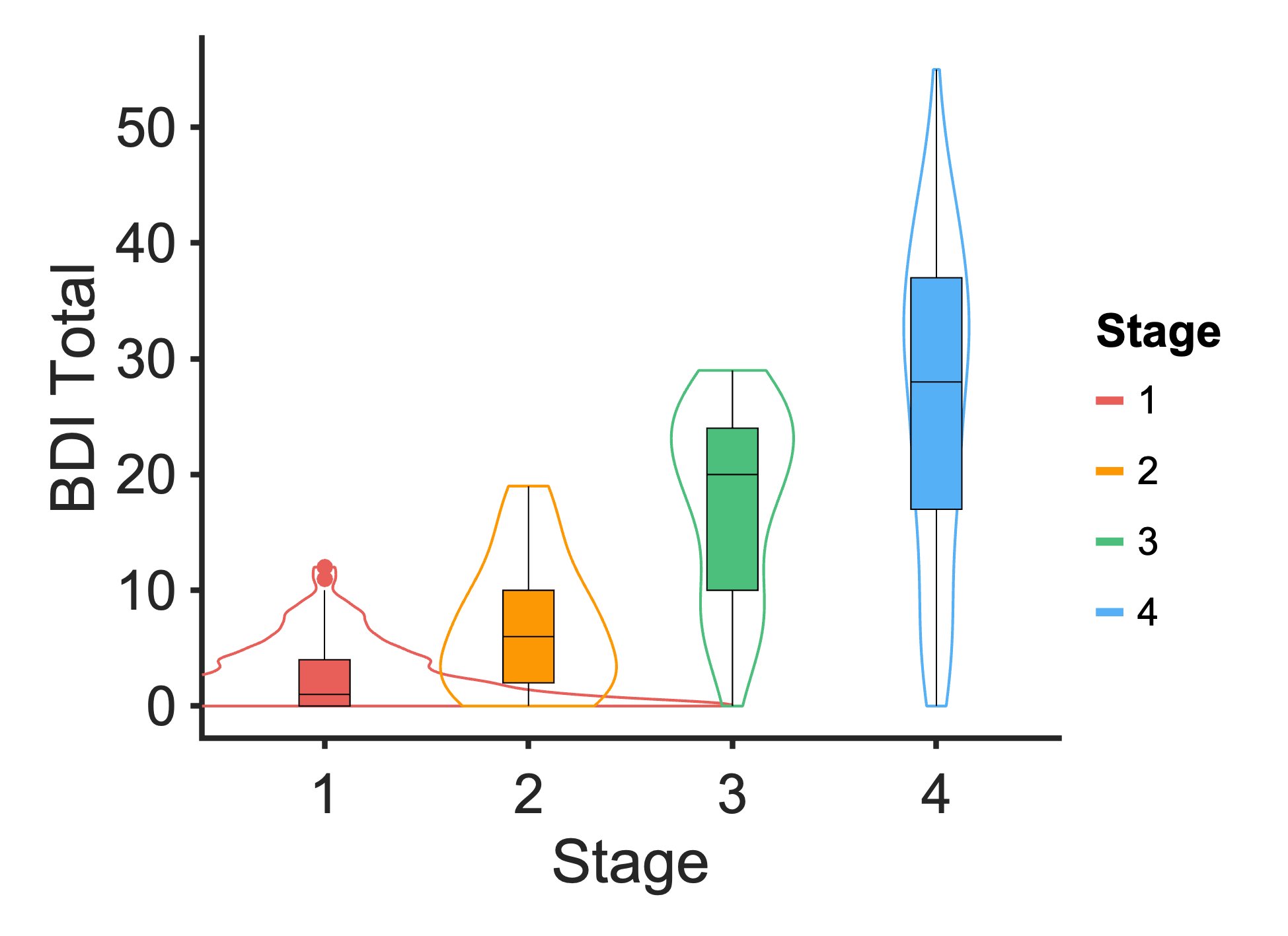

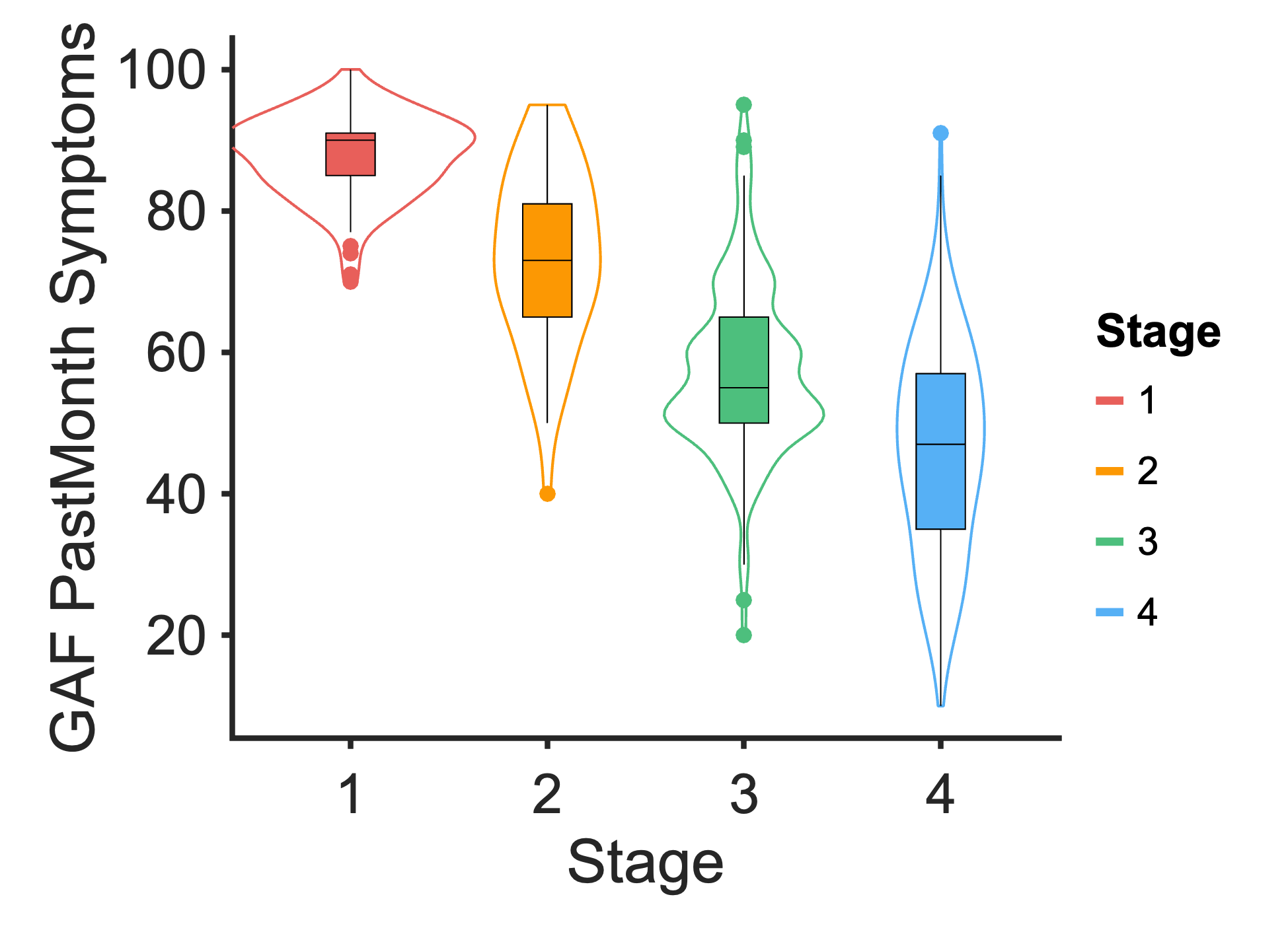

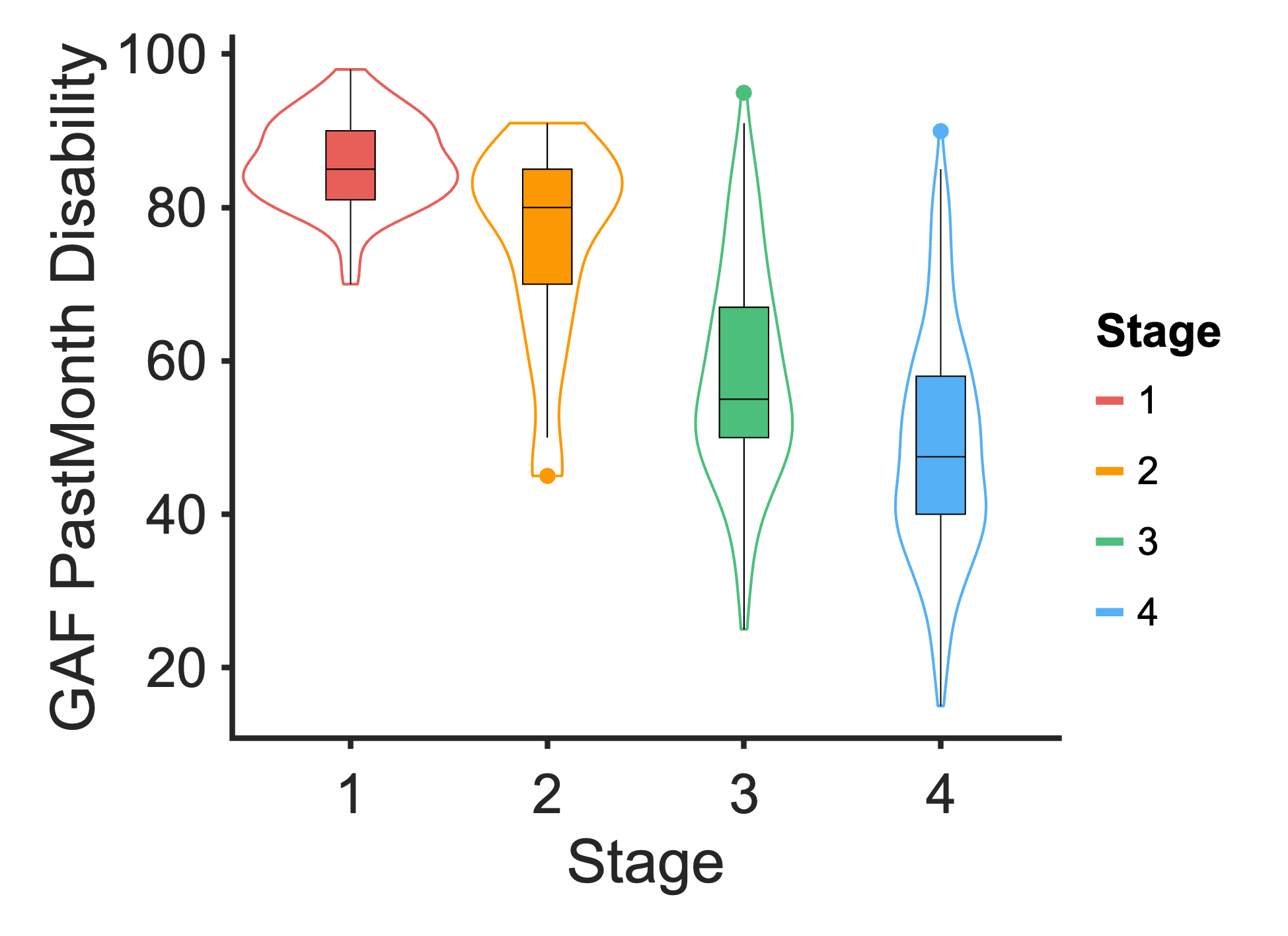

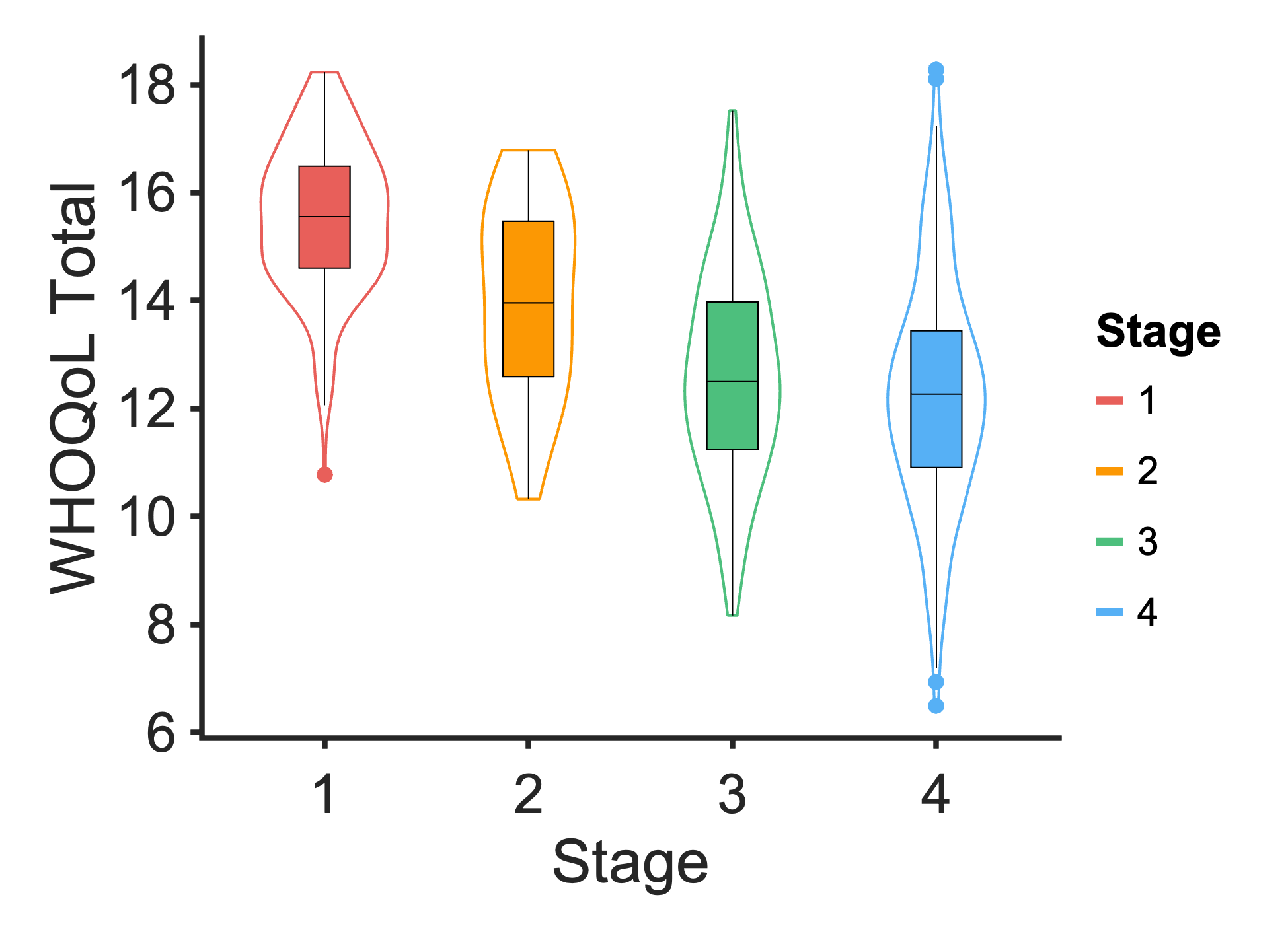

No symptoms	Mild	Moderate	Severe

No symptoms	Mild	Moderate	Severe

None	Mild	Moderate	Severe

No symptoms	Mild	Moderate	Severe

None	Mild	Moderate	Severe

Symptom severity

Symptom severity

Symptom severity

Severity Group

Severity Group

No symptoms	Mild	Moderate	Severe

No symptoms	Mild	Moderate	Severe

No symptoms	Mild	Moderate	Severe

Symptom severity

Symptom severity

Symptom severity

Severity Group

Severity Group

Severity Group

None	Mild	Moderate	Severe

None	Mild	Moderate	Severe

None	Mild	Moderate	Severe

### Figure S2: Illness course of symptom severity groups over 18 months in the discovery sample.

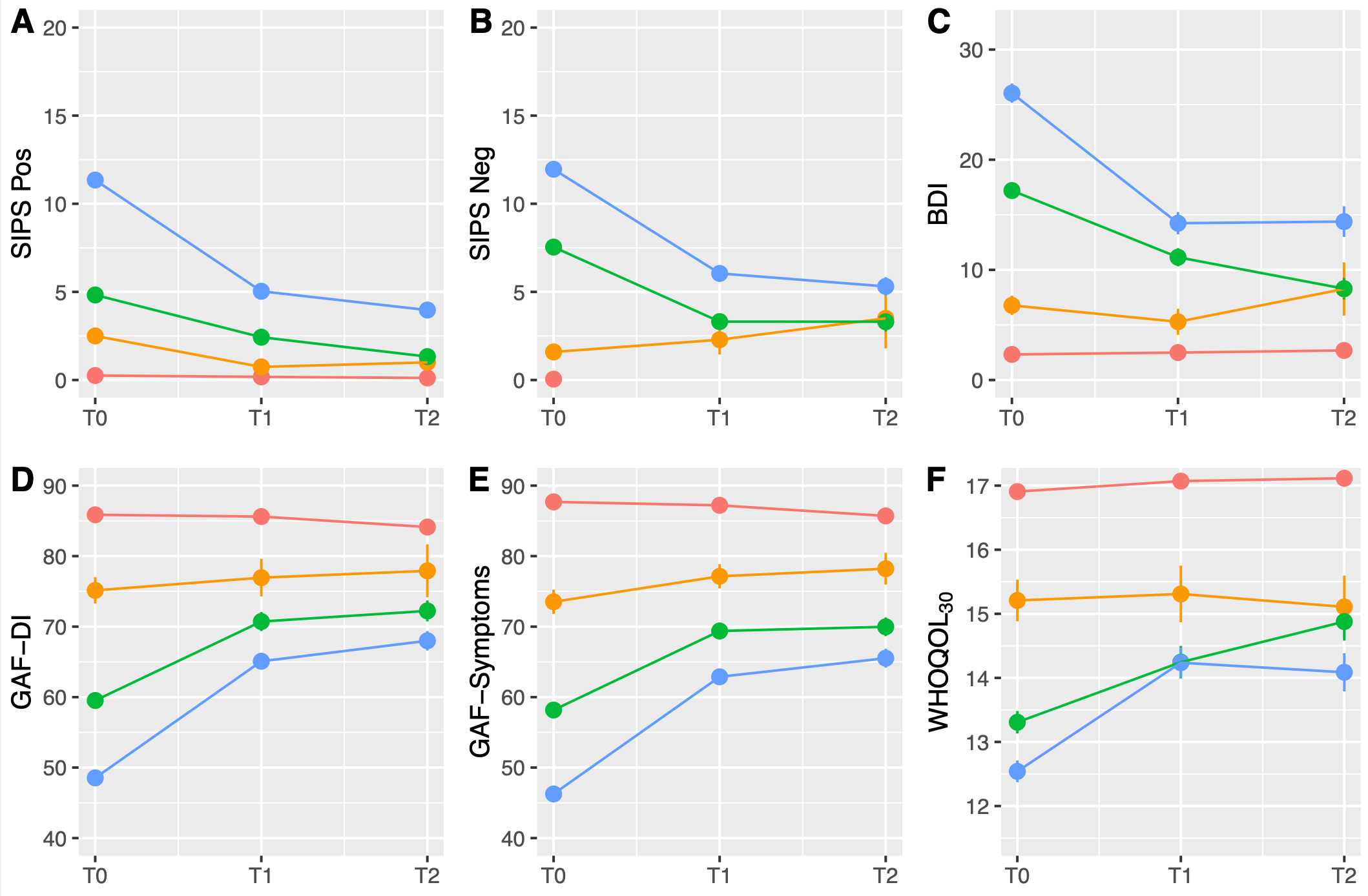

No symptoms
Mild
Moderate
Severe

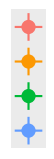

Notes: Lines represent the fitted predicted values and dots are observed mean (SE) values, 95% confidence intervals are shown. Overall, higher symptom severity was associated with a worse longitudinal outcome trajectory. SIPS negative was not measured longitudinally for HC.

### Figure S3: Comparison of illness course of symptom severity groups over 18 months between the original discovery sample and combined discovery and replication sample, controlling for CHR-P and ROD with ROP removed.

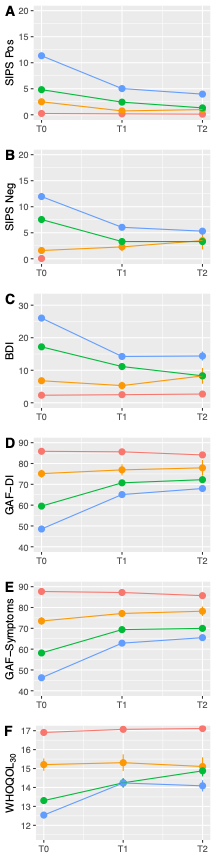

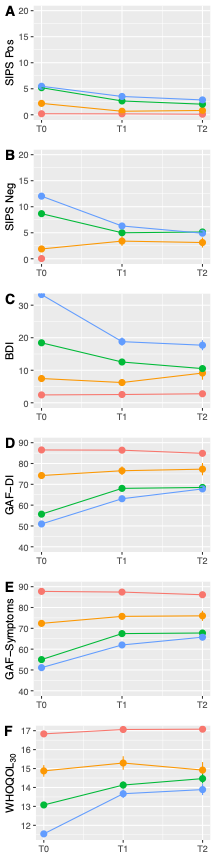

No symptoms
Mild
Moderate
Severe

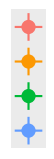

Notes: Lines represent the fitted predicted values and dots are observed mean (SE) values. Longitudinal trajectories were largely the same after controlling for ROD and CHR diagnostic groups.

### Figure S4: SCZ-PRS across symptom severity groups in combined discovery sample without ROP, controlling for CHR-P and ROD

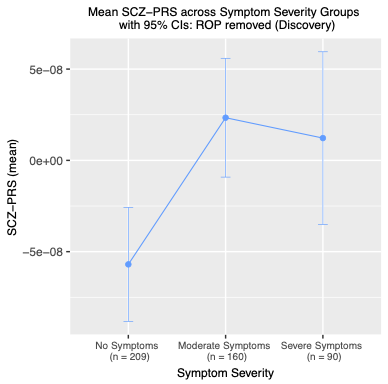

### Note: The mild group was removed from the main analyses due to small group size (n = 43)

### Figure S5: PRS across symptom severity groups with the Mild group included.

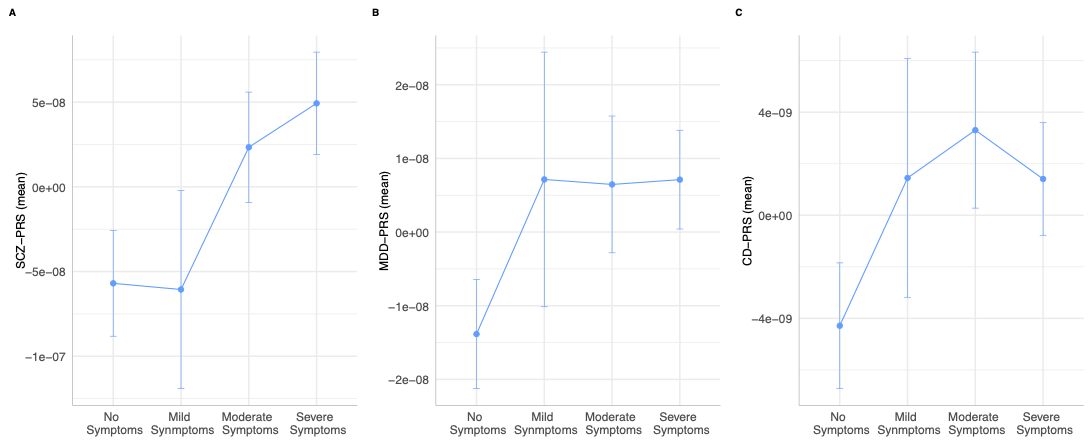

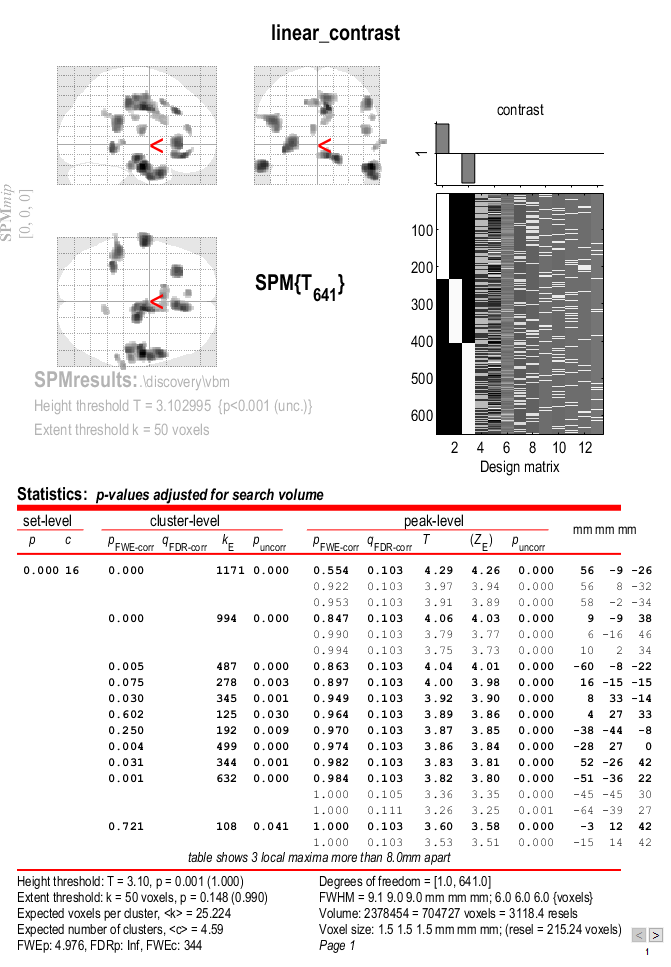

### Figure S6: Discovery *t*-statistical maps showing areas of GMV differences across symptom severity groups using a linear contrast (*p* < .001, uncorr.).

Notes: GMV = Grey Matter Volume; Clusters are labelled in Table S8.

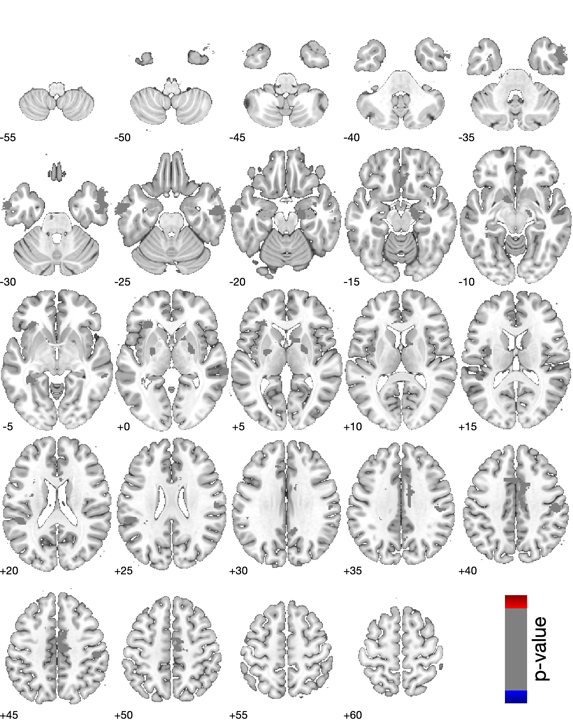
Figure S7: Whole brain plots showing linear GMV differences across symptom severity groups in the discovery sample.

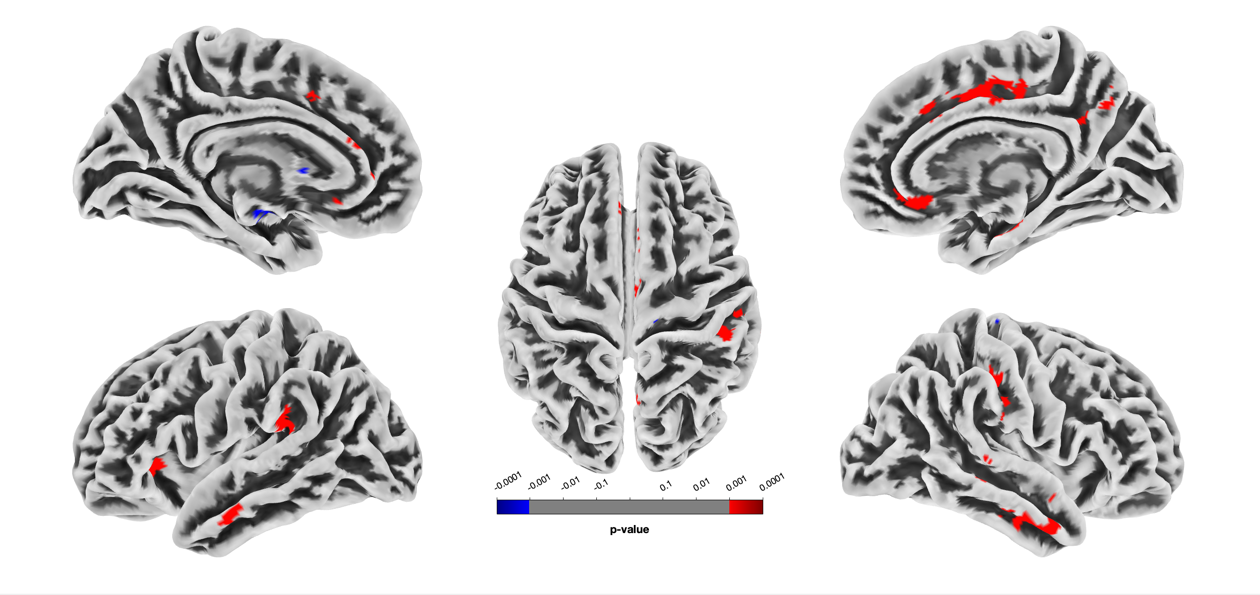

Notes: TIV-corrected surface and subcortical volumes comparing severity groups showing a liner trend in decreasing GMV. Uncorrected *p* < .001.

### Figure S8: Discovery *t*-statistical maps showing areas of GMV differences across symptom severity groups including the Mild group using a linear contrast (*p* < .001, uncorr.).

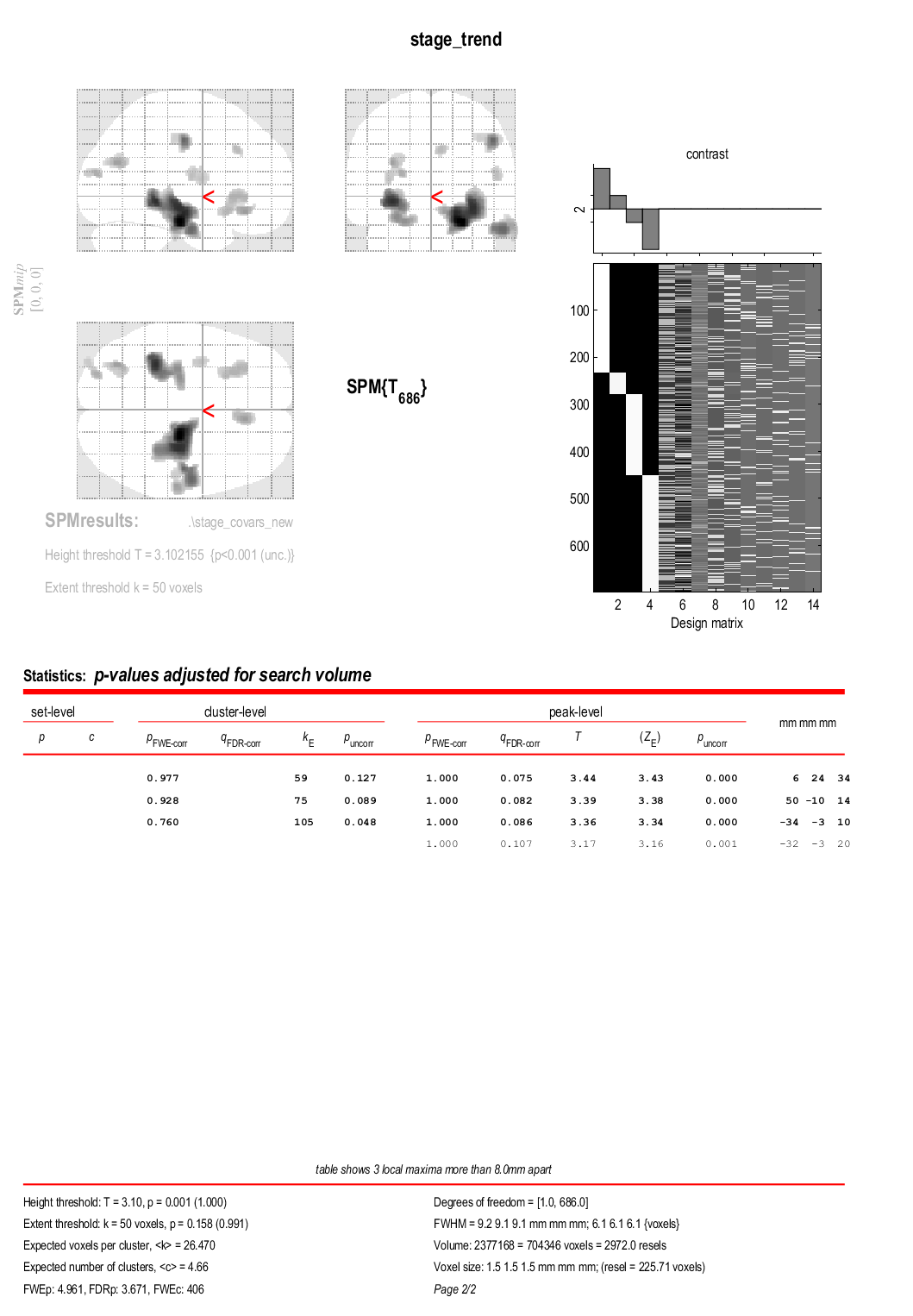

Notes: GMV = Grey Matter Volume. Clusters labelled in Table S10

### Figure S9: Symptom severity group comparison on clinical measures of depressive, positive, and negative symptoms, self-reported functioning, and clinician-reported quality of life in the replication sample

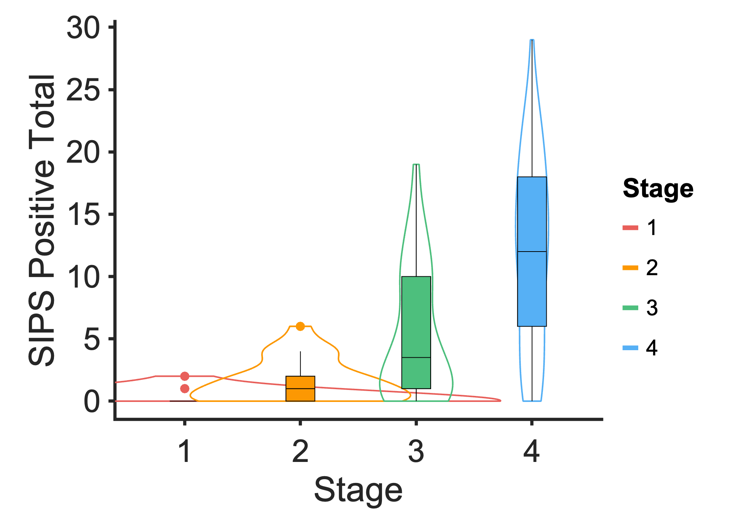

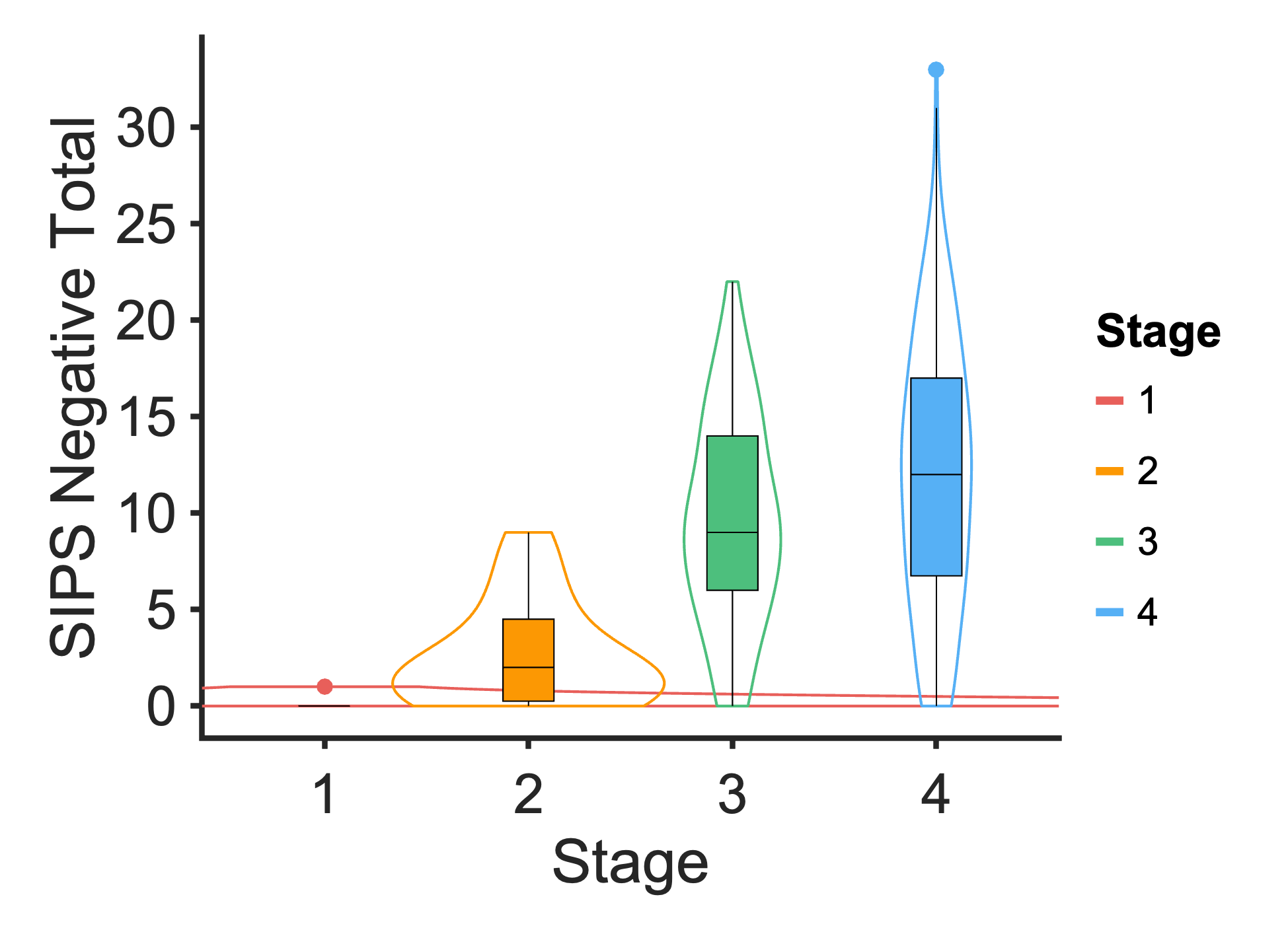

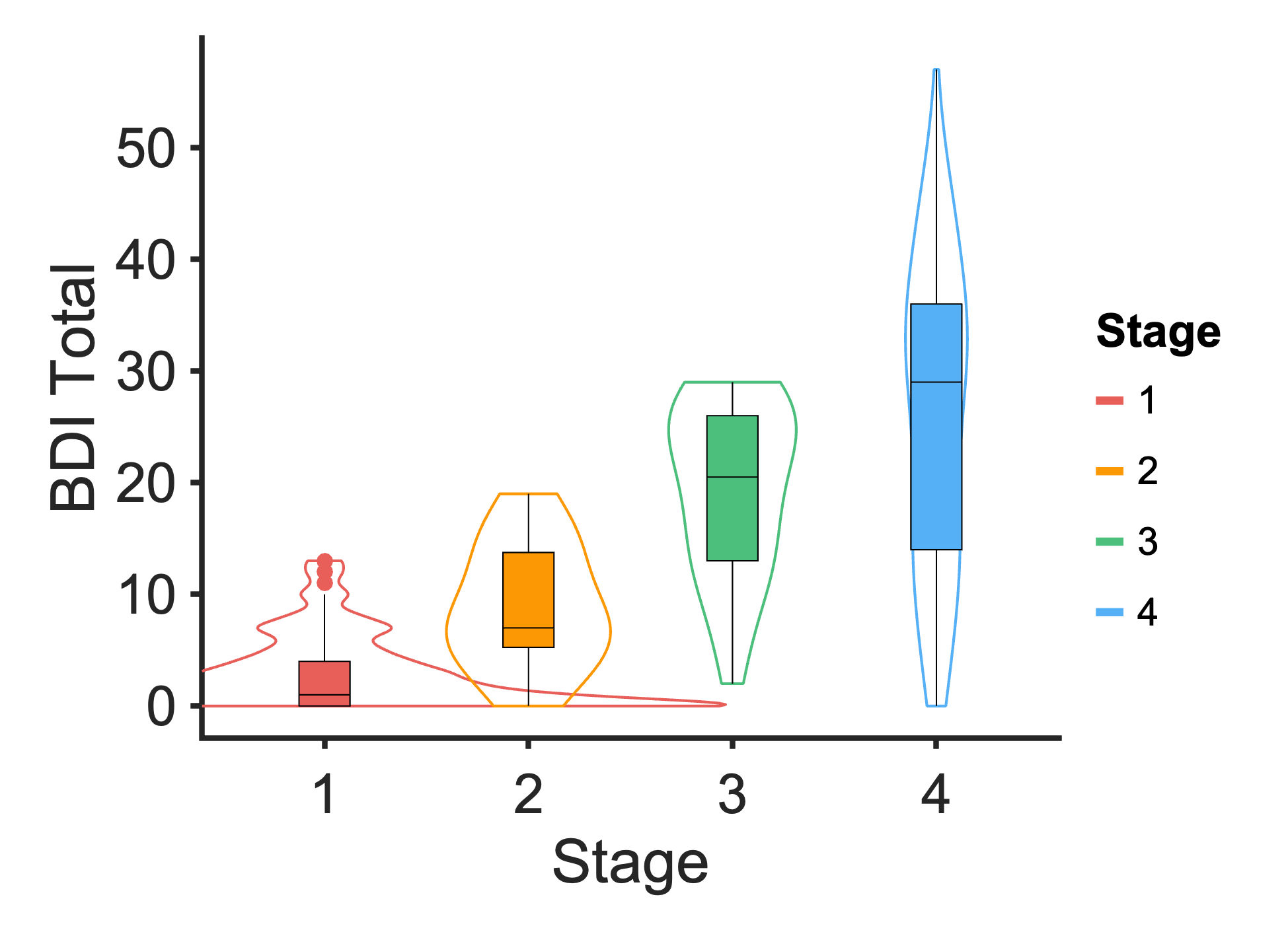

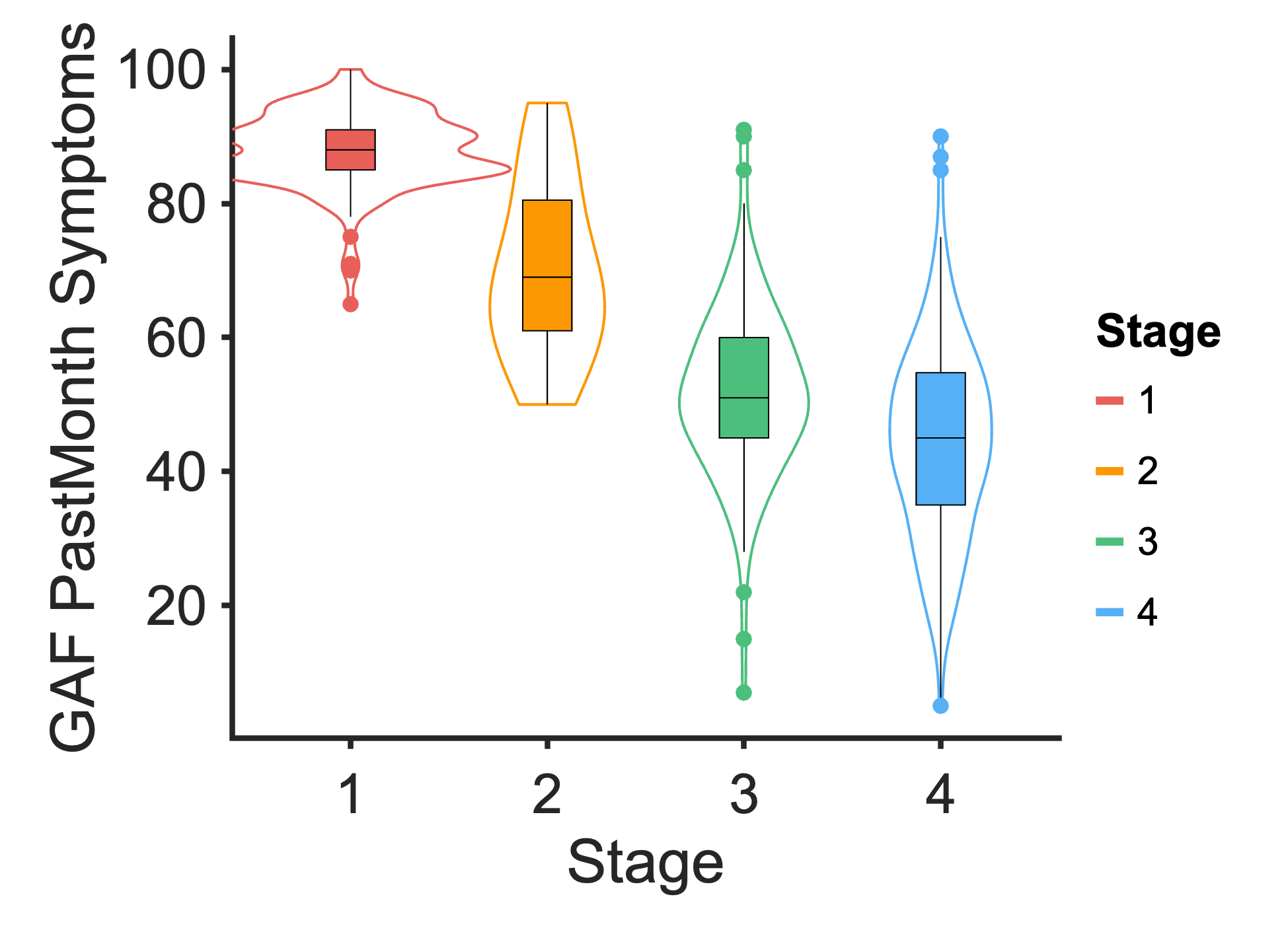

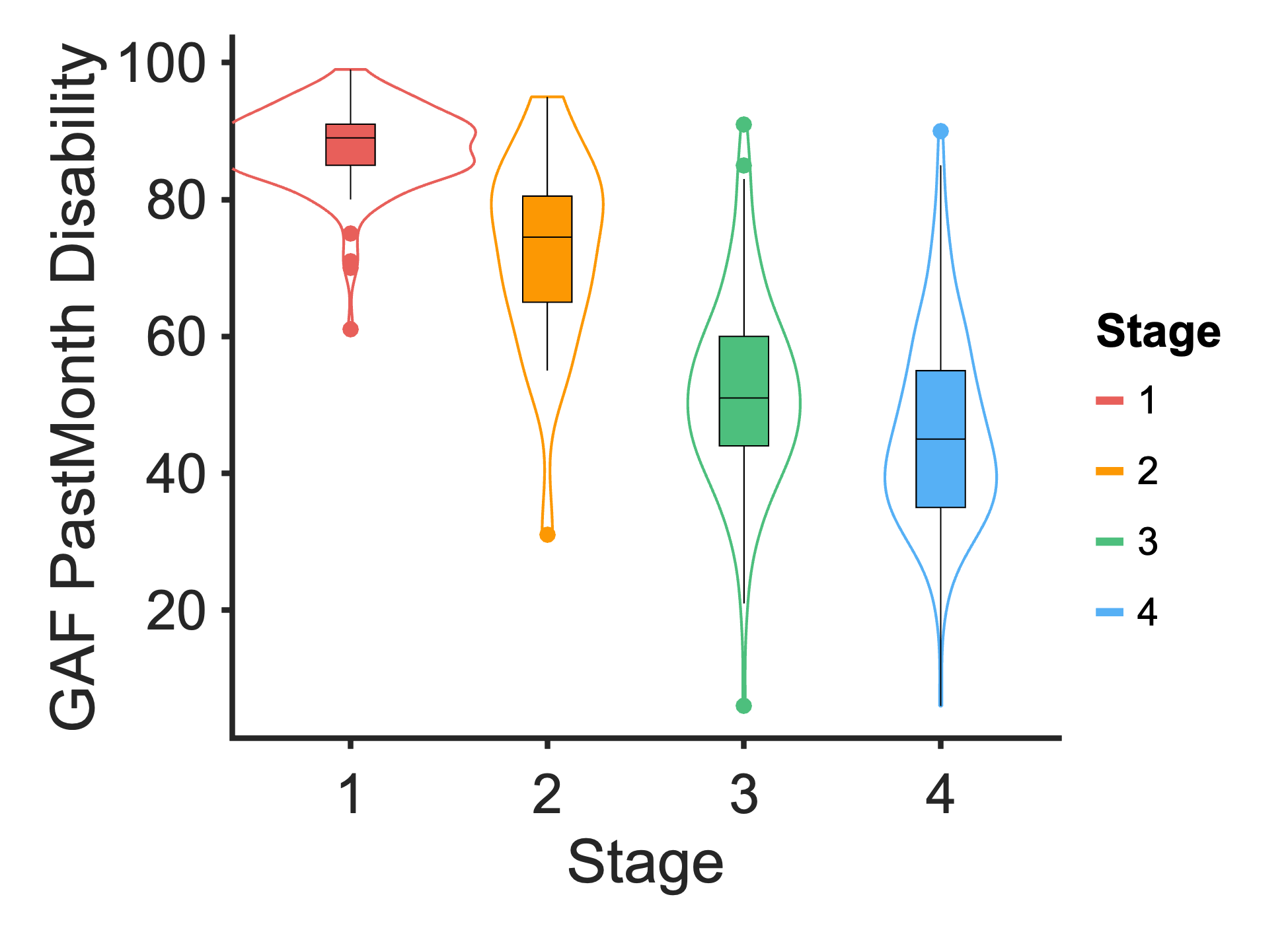

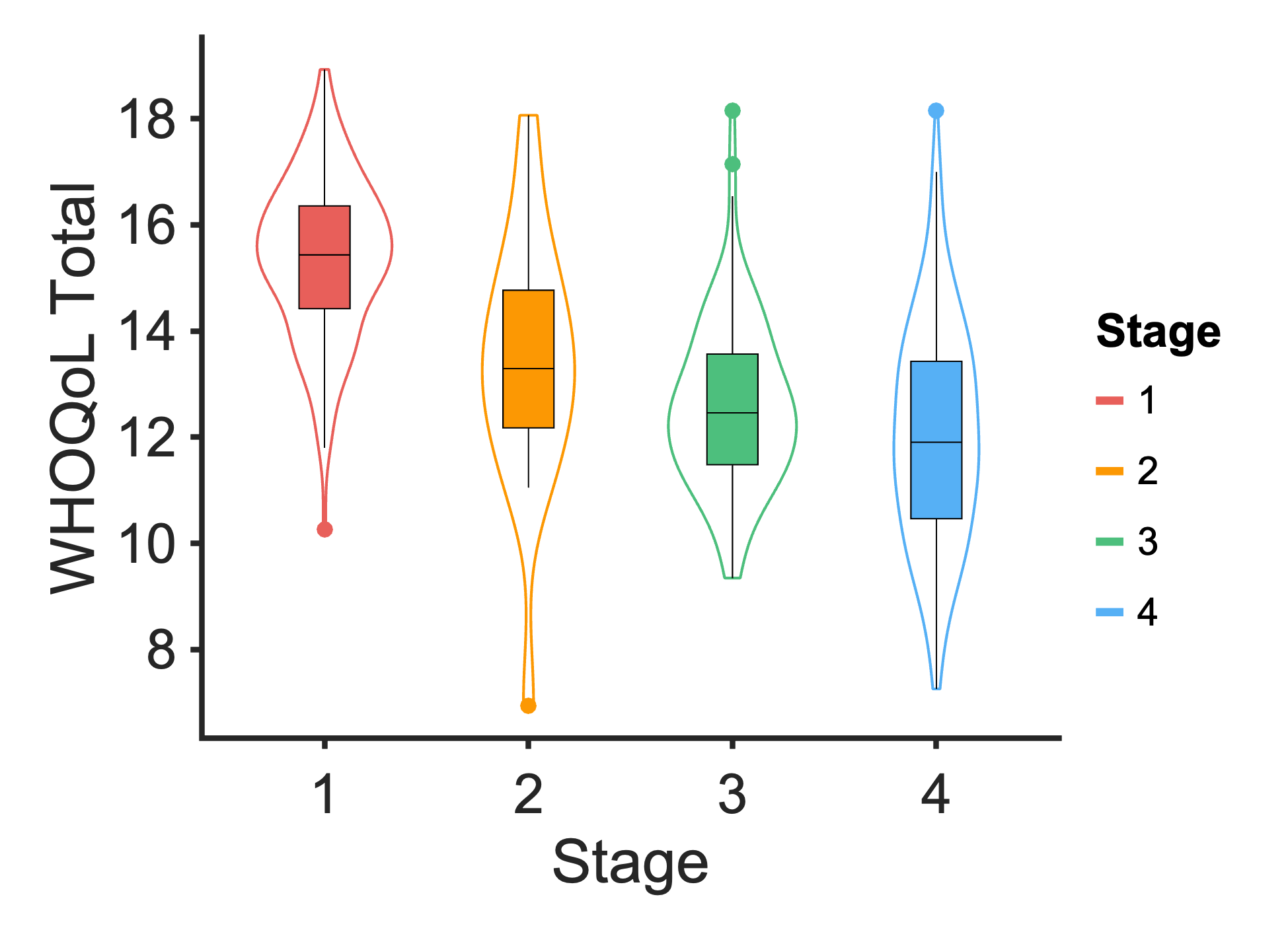

No symptoms	Mild	Moderate	Severe

Symptom Severity

No symptoms	Mild	Moderate	Severe

No symptoms	Mild	Moderate	Severe

No symptoms	Mild	Moderate	Severe

No symptoms	Mild	Moderate	Severe

No symptoms	Mild	Moderate	Severe

Symptom Severity

Symptom Severity

Symptom Severity

Symptom Severity

Symptom Severity

### Figure S10: Comparison of linear mixed models of the discovery and replication samples

No symptoms
Mild
Moderate
Severe

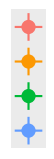

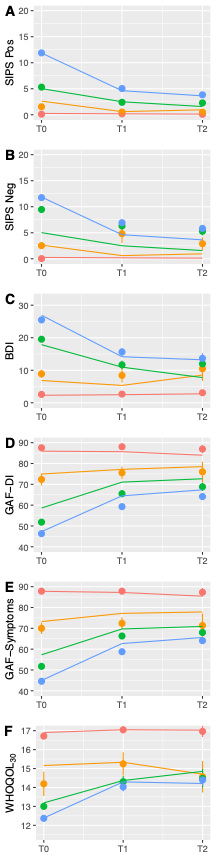

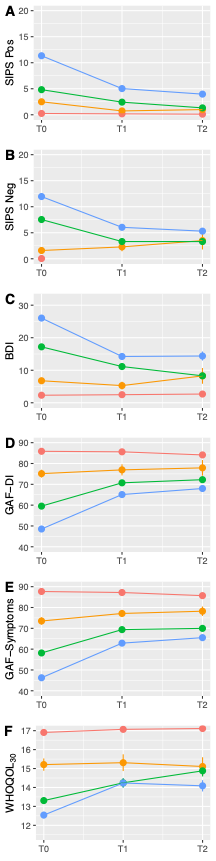

Notes: Linear mixed models from the discovery sample were applied without modification to the replication sample. Lines represent the fitted predicted values and the dots are SE values. Patterns in the discovery sample are mostly replicated in the replication sample. On average, individuals in the replication sample have higher negative symptoms, which is likely a representation of the general baseline differences in clinical symptoms of the sample compared to the discovery sample. Individuals in the replication sample also appear to experience poorer functioning in the symptoms domain across symptom severity groups.

### Figure S11: Differences in premorbid functioning from 0-18 years across symptom severity groups in the replication sample

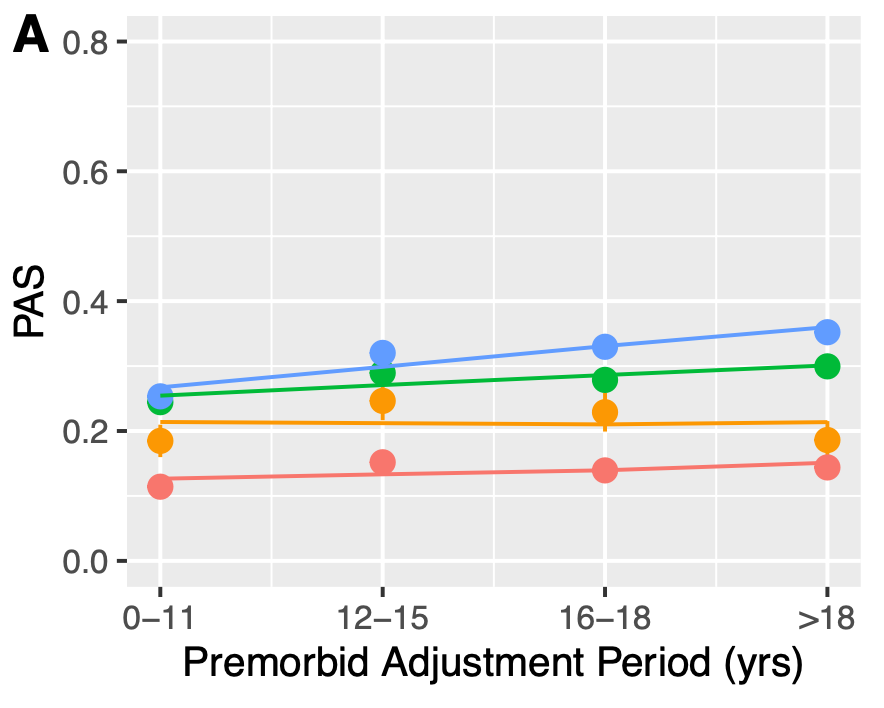

No symptoms
Mild
Moderate
Severe

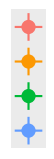

Notes: Linear mixed models from the discovery sample were applied without modification to the replication sample. PAS scores were reversed such that a higher score reflects poorer premorbid functioning.

### Figure S12: Differences in PRS across symptom severity groups in the replication sample (n = 513)

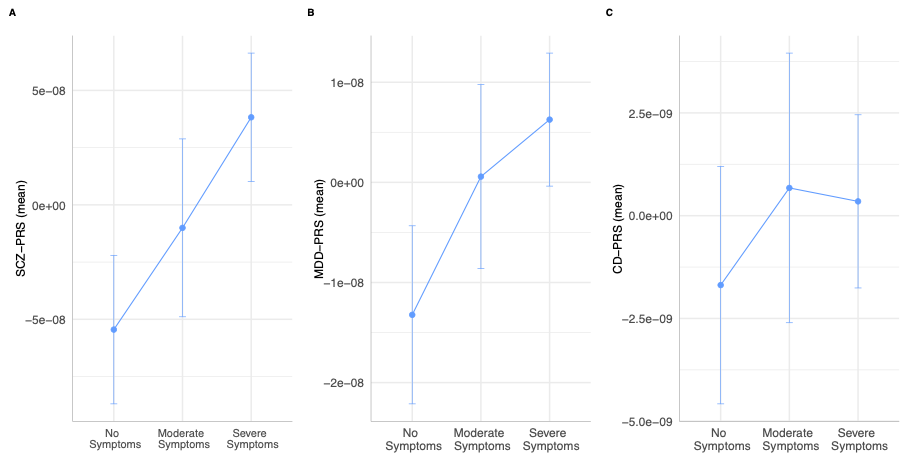

Notes: The mild symptom group was removed due to low sample size (n = 18).

### Figure S13: Replication *t*-statistical maps showing areas of GMV differences across symptom severity groups using a linear contrast (*p* < .001, uncorr.)

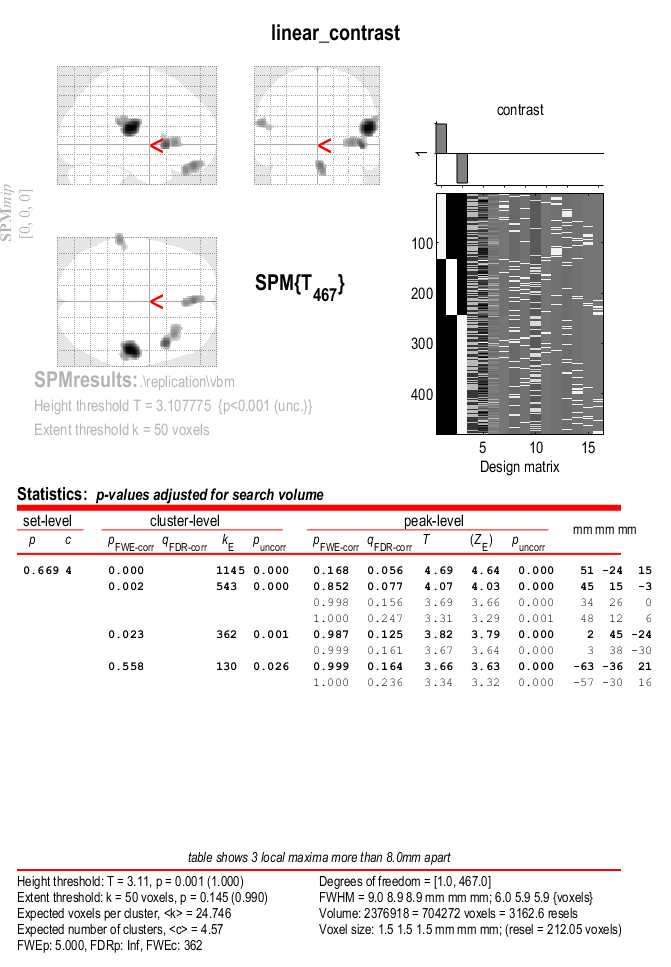

Notes: Clusters are labelled in Table S16.

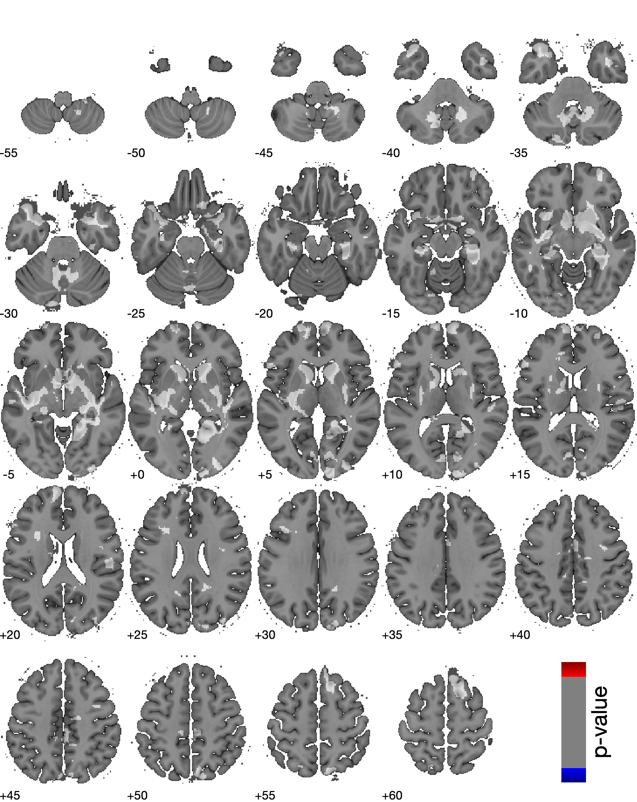
Figure S14: Whole brain plots showing linear GMV differences across symptom severity groups in the replication sample.

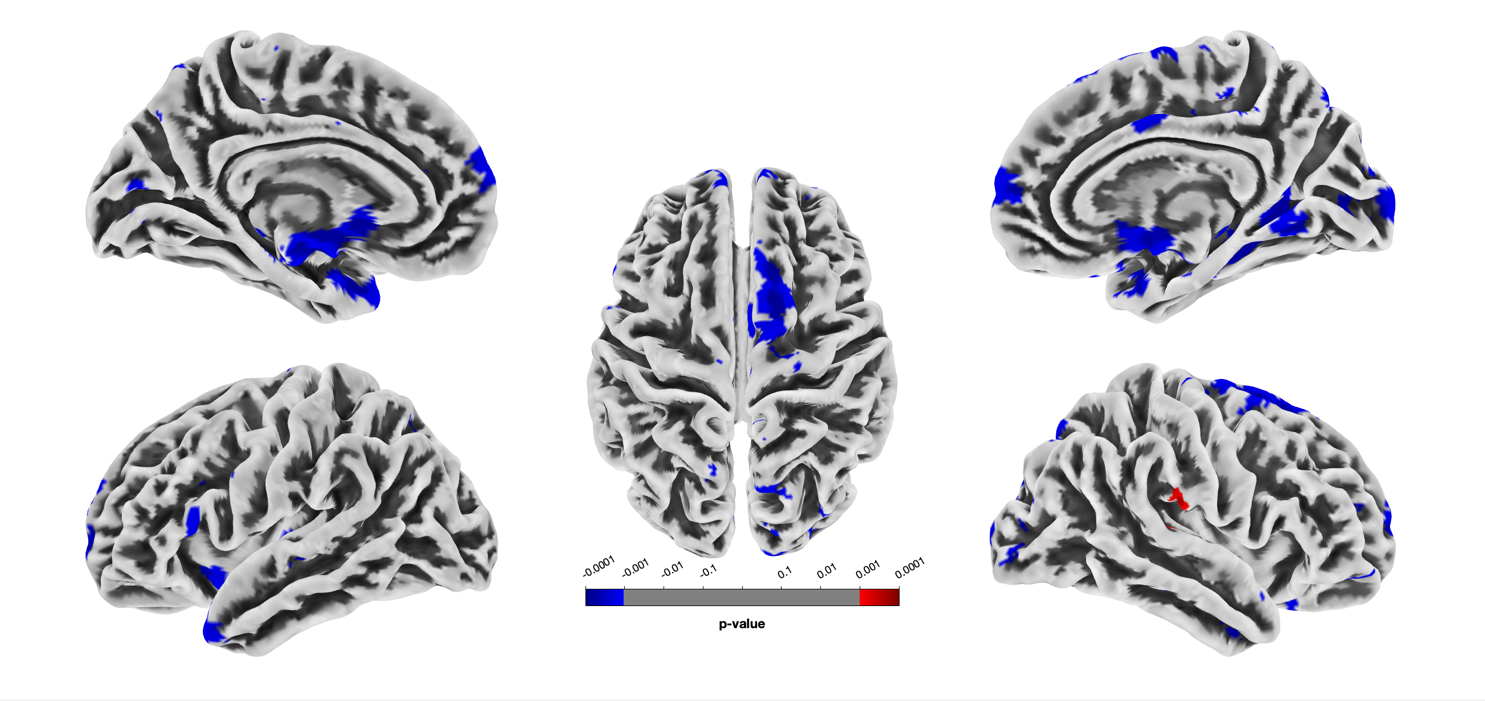

**Supplementary Analyses**

### S1: Brain MRI parameters and processing and analyses

Within PRONIA, a minimal harmonization procedure was used to represent the MRI scanner heterogeneity found in the clinical real-world. Criteria for structural brain imaging were to: 1) Acquire isotropic or nearly isotropic voxel sizes of 1mm resolution; 2) a Field Of View (FOV) to guarantee the full 3D coverage of the brain; 3) the relaxation time (TR) and echo time (TE) to maximise the contrast between cortical ribbon and white matter and to enhance the signal-to-noise ratio in the images. Imaging parameters for each site are included in Table 2. At each site, brain MRI images were visually inspected, automatically defaced, and anonymized using an in-house Freesurfer-based script prior to data centralization. The opensource CAT12 toolbox (version r1205; <http://dbm.neuro.uni-jena.de/cat12/>)^4^ was used to segment images into grey matter (GM), white matter (WM), and cerebrospinal fluid (CSF) maps, and then to high-dimensionally register them to the stereotactic space of the Montreal Neurological Institute (MNI-152 space). The standard processing pipeline of the CAT12 toolbox was used including a) denoising using Spatially Adaptive Non-Local Means filtering; b) Adaptive Maximum A Posteriori (AMAP) segmentation; c) Markov Random Field denoising; d) Local Adaptive Segmentation (LAS) that adjusts the images for inhomogeneity; e) partial volume segmentation; f) high-dimensional DARTEL registration to the MNI-template. The registered GM, WM, and CSF are multiplied by the Jacobian determinants to produce volume maps, and these are segmented into regional volumes using the Hammer’s atlas that consists of 204 regions. Mean GM, WM, CSF, and whole-brain volume (WBV) were compared using parametric tests (FDR, p <0.05). Whole-brain comparisons were conducted with modulated and proportionately scaled GM and WM volume maps smoothed at a full-width at half maximum (FWHM) Gaussian kernel of 8mm. The Quality Assurance framework of CAT12 was used to quantitatively assess the quality of the brain maps. This procedure produces a composite image quality rating (IQR) score that combines measures of the noise contrast ratio, the inhomogeneity contrast ratio, and the image resolution ^4^. Participants were excluded if the IQR was rated as grade D “Sufficient” (n = 4). Brain volumes were proportionately scaled to the total intracranial volume (TIV) of each individual in order to account for global differences in head size. Preprocessing steps during VBM analyses controlled for age, sex, and IQR using regression.

Whole brain VBM regression was performed using SPM12 running in MATLAB (R2024a) to compare GMV between severity groups. Statistical t-maps were enhanced using threshold-free cluster enhancement (TFCE) as it is more sensitive than traditional cluster-based statistical methods to finding true signal than voxel-wise thresholding and reduces false positives^5^(<https://www.neuro.uni-jena.de/tfce/>). One-sided *p-*values were assessed at a statistical significance level of FDR-corrected *P* < .05 for multiple comparisons.

To assess replication of brain findings, region-wise correlation of the discovery and replication brain statistical maps was performed. This process included parcellating the statistical maps using Schaefer cortical and Tian subcortical (https://github.com/yetianmed/subcortex/tree/master/Group-Parcellation/7T) atlases. Parcellation was applied to extract region-wise (parcel-level) data and the two parcel-level maps were correlated. Null distributions for significance testing was generated using a spin-test approach to randomise data while preserving spatial relationships^6^.

### S2: Subject and feature filtering

A total of 22 participants were removed from analyses due to missing more than 50% of baseline data. The missing cases also did not have any MRI data. The 22 participants removed did not differe significantly from the rest of the sample on clinical and functioning variables (Table S2). From the baseline feature list, features were excluded if they had more than 95% of the same values or if more than 50% of participants were missing data.

### Premorbid and longitudinal analyses

Premorbid functioning was assessed with the premorbid adjustment scale (PAS), which determines premorbid psychosocial functioning by assessing the attainment of developmental milestones in four categories: social accessibility-isolation, peer relationships, ability to function outside the family, and capacity to form intimate sociosexual relationships. These four categories are assessed across four life periods: childhood (<12 years), early adolescence (12-15 years), late adolescence (16-18), and adulthood (>18 years). A higher score indicates poorer premorbid functioning.

Longitudinal outcome measures were chosen based on their importance for prognoses in terms of symptoms and functional outcomes, making them suitable measures for illness severity. Further, the chosen measures are easily interpretable as prognostic markers and can be analysed with conventional inferential statistics. Clinical measures include BDI depression, SIPS positive symptoms, SIPS negative symptoms, GAF disability scale, GAF symptoms scale, and WHOQoL total. Total scores were used for each measure to increase interpretability of the results and to minimise the amount of multiple comparisons to avoid false positives. Data from baseline, 9-month (T1), and 18-month (T2) follow-up were extracted for each measure.

Premorbid functioning and longitudinal illness trajectories were investigated using mixed models in the discovery sample (R package *lme4*; Satterthwaite approximation of degrees of freedom, cases and intercepts modelled as random effects). Main fixed effects of severity groups and time, linear trends, quadratic trends, and trend interactions with severity group were assessed. Post-hoc tests (EMMEANS) were used to compare trends of each measure between severity groups (false-discovery rate corrected for multiple comparisons, significant at p < .05). Effect sizes for each complete model (marginal R2) was calculated according to Nakagawa and Schielzeth ^7^ and Johnson ^8^ as implemented in R package *r2glmm*^9^.

### Table S17: Kruskal-Wallis tests for baseline clinical variables across symptom severity groups in the discovery sample

| Variable | No symptoms | Mild | Moderate | Severe | F/Chi2(SD) | Sig | Eta2/Phi |
| --- | --- | --- | --- | --- | --- | --- | --- |
| N | 241 | 50 | 182 | 254 |  |  |  |
| Clinical High Risk, N(%) | 0(0.0) | 0(0.0) | 94(51.6) | 49(19.3) | 189.07(8) | 9.70e-41 | 0.51 |
| Recent Onset Depression, N(%) | 0(0.0) | 21(42.0) | 76(41.8) | 54(21.3) | 125.63(8) | 4.74e-27 | 0.42 |
| Recent Onset Psychosis, N(%) | 0(0.0) | 0(0.0) | 0(0.0) | 150(59.1) | 351.95(8) | 5.65e-76 | 0.70 |
| Control Participants, N(%) | 241(100.0) | 29(58.0) | 12(6.6) | 1(0.4) | 624.43(8) | 5.09e-135 | 0.93 |
| Age, yrs(SD) | 25.8(6.4) | 25.5(6.5) | 24.7(5.6) | 25.3(5.7) | 1.66(3,723) | -- | -- |
| Gender, male(%) | 93(38.6) | 20(40.0) | 97(53.3) | 134(52.8) | 14.06(8) | 2.82e-03 | 0.14 |
| Education, yrs(SD)) | 15.9(3.0) | 15.1(3.1) | 14.1(3.1) | 14.1(2.9) | 49.12(3,717) | 1.23e-10 | 0.07 |
| Marital status, single(%) | 114(47.5) | 32(66.7) | 133(73.1) | 184(72.7) | 56.55(20) | 9.53e-08 | 0.28 |
| Employment status, unemployed(%) | 1(0.4) | 0(0.0) | 18(9.9) | 52(20.5) | 62.37(8) | 1.83e-13 | 0.29 |
| Family psychosis risk, yes(%) | 5(2.1) | 2(4.0) | 19(10.4) | 29(11.4) | 58.24(36) | 1.13e-04 | 0.28 |
| Birth complications, yes(%) | 28(11.7) | 6(12.0) | 24(13.2) | 41(16.2) | 2.35(8) | -- | -- |
| Head Trauma, yes(%) | 21(8.8) | 2(4.0) | 31(17.0) | 57(22.5) | 23.49(8) | 3.20e-05 | 0.18 |
| SIPS Positive, mean(SD) | 0.3(0.6) | 2.5(2.2) | 5.2(4.4) | 11.4(7.5) | 425.45(3,719) | 6.78e-92 | 0.47 |
| SIPS Negative, mean(SD) | 0.1(0.2) | 1.6(2.1) | 8.1(5.6) | 12.0(7.0) | 495.19(3,716) | 5.27e-107 | 0.51 |
| SIPS Disorganised, mean(SD) | 0.1(0.3) | 0.7(1.2) | 2.4(2.1) | 4.5(3.8) | 370.51(3,716) | 5.40e-80 | 0.36 |
| SIPS General, mean(SD) | 0.3(0.8) | 2.4(2.8) | 6.4(3.4) | 8.1(4.2) | 436.33(3,716) | 2.98e-94 | 0.55 |
| SIPS Total, mean(SD) | 0.6(1.2) | 7.1(4.6) | 22.1(10.3) | 36.0(15.4) | 555.94(3,713) | 3.59e-120 | 0.67 |
| SANS Total, mean(SD) | NaN(NaN) | 10.1(10.1) | 18.6(16.8) | 29.2(24.8) | 25.58(2,374) | 2.79e-06 | NaN |
| BDI Total, mean(SD) | 2.3(2.8) | 6.8(5.6) | 17.8(8.0) | 26.3(13.4) | 411.26(3,668) | 8.07e-89 | 0.56 |
| GAF social past month, mean(SD) | 87.7(5.6) | 73.5(12.3) | 57.2(12.1) | 46.1(14.5) | 507.03(3,722) | 1.43e-109 | 0.71 |
| GAF disability past month, mean(SD) | 85.9(5.5) | 75.1(13.2) | 58.3(13.8) | 48.5(14.6) | 472.07(3,722) | 5.39e-102 | 0.64 |
| WHO QoL Total | 15.5(1.4) | 14.0(1.7) | 12.6(1.9) | 12.3(2.1) | 269.29(3,630) | 4.40e-58 | 0.40 |
| WAIS-V Vocabulary | 11.9(2.8) | 11.4(3.0) | 10.8(2.9) | 10.4(3.4) | 29.08(3,700) | 2.15e-06 | 0.04 |
| WAIS-V Matrix Reasoning | 11.2(2.2) | 11.2(2.6) | 10.7(2.6) | 10.1(2.7) | 18.46(3,652) | 3.54e-04 | 0.04 |

### Table S19: Reclassification of original PRONIA study groups into symptom severity groups

|  | Positive | Negative | Depression | Positive & Negative | Positive & Depression | Negative & Depression | All symptoms | Total N |
| --- | --- | --- | --- | --- | --- | --- | --- | --- |
| Severe symptoms | | | | | | | | |
| HC | 0 | 0 | 1 | 0 | 0 | 0 | 0 | 1 |
| ROD | 0 | 7 | 43 | 0 | 1 | 1 | 0 | 52 |
| CHR-P | 0 | 7 | 35 | 0 | 0 | 6 | 0 | 48 |
| ROP | 89 | 0 | 0 | 31 | 19 | 0 | 11 | 150 |
| Moderate symptoms | | | | | | | | |
| HC | 0 | 7 | 5 | 0 | 0 | 0 | 0 | 12 |
| ROD | 2 | 42 | 5 | 0 | 0 | 29 | 0 | 78 |
| CHR-P | 23 | 0 | 0 | 25 | 13 | 0 | 34 | 95 |
| Mild symptoms | | | | | | | | |

| HC | 21 | 6 | 2 | 0 | 0 | 0 | 0 | 29 |
| --- | --- | --- | --- | --- | --- | --- | --- | --- |
| ROD | 6 | 7 | 3 | 3 | 0 | 2 | 0 | 21 |
| **No symptoms** | | | | | | | | |
| HC |  |  |  |  |  |  |  | 241 |

Notes: This table shows how individuals in the original HC, ROD, CHR-P, and ROP groups were redistributed into mild, moderate, and severe symptom groups based on highest symptom severity. As shown, individuals in CHR-P and ROD had severe negative and depression symptoms that were not captured in their original classifications. A low number of individuals from those study groups also had severe presentations of positive symptoms that id not meet criteria for a psychotic episode (i.e brief, one-dimension positive symptom).

### Table S20: Distribution of CHR-P, ROD, ROP, and HC groups into groups based on positive, negative, and depression symptom severity in the discovery sample

|  | **No symptoms, n (%)** | | | | | | **Mild symptoms, n (%)** | | | | | | **Moderate symptoms, n (%)** | | | | | | **Severe symptoms, n (%)** | | | | |
| --- | --- | --- | --- | --- | --- | --- | --- | --- | --- | --- | --- | --- | --- | --- | --- | --- | --- | --- | --- | --- | --- | --- | --- |
|  | | HC | CHR-P | ROD | ROP | *N* | | HC | CHR-P | ROD | ROP | *N* | HC | CHR-P | ROD | ROP | *N* | HC | | CHR-P | ROD | ROP | *N* |
| **Positive** | | 259 | 0 | 0 | 0 |  | | 24 | 6 | 70 | 0 |  | 0 | 137 | 0 | 0 |  | 0 | | 0 | 0 | 150 |  |
| **Negative** | | 269 | 0 | 0 | 0 |  | | 7 | 30 | 25 | 25 |  | 7 | 89 | 104 | 86 |  | 0 | | 13 | 8 | 42 |  |
| **Depression** | | 274 | 0 | 0 | 0 |  | | 3 | 11 | 23 | 18 |  | 5 | 50 | 38 | 42 |  | 1 | | 41 | 45 | 30 |  |
| **COGDIS** | | 283 | 0 | 0 | 0 |  | | 0 | 0 | 0 | 0 |  | 0 | 6 | 0 | 0 |  | 0 | | 0 | 0 | 0 |  |
| Final assignment | | 241 (85) | 0  (0) | 0  (0) | 0  (0) | 241 | | 29  (10) | 0  (0) | 21 (14) | 0  (0) | 50 | 12  (4) | 94  (66) | 76  (50) | 0  (0) | 182 | 1  (0.3) | | 49  (34) | 54  (36) | 150  (150) | 254 |

Note: Individuals were classified based on the highest symptom.

### Table S21: Comparison of symptom severity group distribution across sites.

| **Variable** | **None** | **Mild** | **Moderate** | **Severe** | **F/Chi2** | ***P*** | **Eta2/Phi** |
| --- | --- | --- | --- | --- | --- | --- | --- |
| N | 241 | 50 | 182 | 254 |  |  |  |
| Basel | 34(14.1) | 4(8.0) | 26(14.3) | 33(13.0) | 1.52(8) | -- | -- |
| Birmingham | 32(13.3) | 10(20.0) | 16(8.8) | 25(9.8) | 6.33(8) | -- | -- |
| Cologne | 60(24.9) | 4(8.0) | 32(17.6) | 42(16.5) | 10.62(8) | 1.39e-02 | 0.12 |
| Milan | 11(4.6) | 4(8.0) | 9(4.9) | 18(7.1) | 2.14(8) | -- | -- |
| Munich | 49(20.3) | 7(14.0) | 49(26.9) | 87(34.3) | 16.60(8) | 8.52e-04 | 0.15 |
| Turku | 26(10.8) | 4(8.0) | 24(13.2) | 34(13.4) | 1.78(8) | -- | -- |
| Udine | 29(12.0) | 17(34.0) | 26(14.3) | 15(5.9) | 32.83(8) | 3.50e-07 | 0.21 |

Notes. F, ANOVA F-score; χ2 , chi-squared value; df, degrees of freedom; p-value, p-value associated with ANOVA or χ2 (only false-discovery rate values shown); Eta2 , eta-squared effect size measure associated with ANOVA tests; Phi, phi coefficient measure of effect size for χ2 tests. ANOVA used for all variables displayed with the mean and standard deviation [mean(SD)] and chi-squared tests used for all nominal variables displayed with the total number and percentage of total [n(%)].

### Table S22: Spearman correlation between symptom dimensions within and across severity groups

|  | BDI | SIPS Positive | SIPS Negative | |
| --- | --- | --- | --- | --- |
| Mild symptoms |  |  | |  |
| BDI | 1.00 | 0.01 | | 0.33 |
| SIPS positive | 0.01 | 1.00 | | 0.17 |
| SIPS negative | 0.33 | 0.17 | | 1.00 |
| Moderate symptoms | | | | |
| BDI | 1.00 | -0.08 | | 0.17 |
| SIPS positive | -0.08 | 1.00 | | 0.22 |
| SIPS negative | 0.17 | 0.22 | | 1.00 |
| Severe symptoms | | | | |
| BDI | 1.00 | -0.47 | | -0.01 |
| SIPS positive | -0.47 | 1.00 | | 0.19 |
| SIPS negative | -0.01 | 0.19 | | 1.00 |
| Across sample |  |  | |  |
| BDI | 1.00 | 0.52 | | 0.68 |
| SIPS positive | 0.65 | 1.00 | | 0.65 |
| SIPS negative | 0.68 | 0.55 | | 1.00 |

### S3: Summary of the distribution of original study groups, symptoms, and within- and across- group correlations

There was a significant difference in distribution of CHR-P, ROD, and ROP across mild (X^2^ = 157, *df* = 2, *P* < .001), moderate (X^2^ = 426, *df* = 2, *P* < .001), and severe (X^2^ = 427, *df* = 2, *P* < .001) positive symptoms, and in severe negative symptoms (X^2^ = 36.4, *df* = 2, *P* < .001). Only 6 individuals met criteria for COGDIS exclusively. These 6 individuals were combined with the moderate positive symptom group for subsequent analyses based on CHR-P representation and the close association between COGDIS and positive symptoms, and psychosis outcome^10,11^.

There was a significant difference in the distribution of positive, negative, and depressive symptoms in the mild (X^2^ = 11.3, *df* = 2, *P* < .01), moderate (X^2^ = 76.8, *df* = 2, *p* < .001), and severe (X^2^ = 37.6, *df* = 2, *P* < .001) groups.

Within severity groups, symptom dimensions showed small Spearman’s correlations, except for a large negative correlation between BDI and SIPS positive scores in the severe group (ρ = -0.47), and a medium positive correlation between BDI and SIPS negative scores in the mild group (ρ = 0.33). Across groups, symptom dimensions were largely correlated.

While CHR-P, ROD, and ROP individuals were distributed across transdiagnostic severity groups, there were differences in how individuals were re-classified. Mild, moderate, and severe depression symptoms were distributed across all study groups, which supports previous research^12,13^. In contrast, positive and negative symptom distribution differed across study groups, likely reflecting recruitment methods and the core role of these symptoms in psychotic disorders^14^. Thus, positive symptoms may be overall more disorder-specific^15.^.^.^ As expected, we observed that CHR-P patients present with more severe symptoms beyond positive symptoms^16^, highlighting the importance of assessing and treating comorbidities as per current guidelines^17^, and monitoring mild or emerging symptoms^15^ that may be masked by more severe clinical presentations.

Symptom dimensions were moderately to strongly correlated across the sample, but only weak to moderate within groups, indicating that higher severity in one symptom dimension (i.e. depression) is not always associated with elevation in others (i.e. positive). Interestingly, while the positive depression-negative symptom correlation weakened with increasing severity, the negative depression-positive correlation increased with increasing severity, possibly reflecting shared severity trajectories of positive and negative symptoms, but distinct severity trajectories when depression is paired with either negative or psychotic symptoms. The strong overall correlations may reflect some shared underlying mechanisms of illness across dimensions in more severe cases and indicate increased co-occurrence. This aligns with literature showing that more severe illness presentations often involve transdiagnostic symptom profiles compared to symptoms at milder severity^18^, emphasising the importance of a transdiagnostic approach to stratifying illness severity.

### S4: Diagnostic effects of severity groups

There may be concerns that diagnosis-specific differences between ROD and CHR-P/ROP have contaminated our groups. To test for this, we evaluated whether the distinction in diagnostic group (ROD, CHR) drove differences in the symptom severity groups. HC was not controlled due to minimal clinical symptoms and overlap with the no symptoms group.

Baseline and longitudinal analyses were rerun without the ROP group, as all ROP individuals had severe positive symptoms, therefore, they would not contribute to variation in the groups. HCs were included in the analyses as a reference group. To increase power, we combined the discovery and replication samples for a combined sample of n = 997. We controlled for ROP and CHR by adding them as covariates in our linear mixed model. The same control analyses were performed for the genetic and brain analyses. At baseline, differences across symptom severity groups were maintained on all measures after removing ROP (Table S7). Longitudinal trends of clinical and functioning measures after controlling for ROD and CHR were comparable to the original findings, indicating that the variations in symptom severity groups were not driven by diagnostic group (Figure S3). SIPS positive scores are notably lower and BDI scores higher in the controlled sample for the severe symptom group due to removal of the ROP diagnostic group. There are some differences in functioning for the severe symptom group between the discovery sample and controlled sample. Differences in functioning may suggest that ROP status influences functioning in the group.

For SCZ-PRS, after removing ROP and controlling for CHR-P and ROD, there were significant differences between no and moderate symptoms (*P* < .01) and no and severe symptoms (*P* < .05) groups, but not between moderate and severe symptoms groups (*P* = .9). The results suggest that the inclusion of ROP drove the increase in SCZ-PRS of the severe symptom group in the original analyses (Figure S4). The importance of ROP inclusion in the SCZ-PRS score of the severe symptom group mirrors recent literature arguing that SCZ-PRS may be more strongly associated with diagnosis (i.e. schizophrenia), and the increase in SCZ-PRS in the moderate symptom group compared to the no symptom group reflects evidence that SCZ-PRS is higher in patients compared to controls, providing useful clinical information regardless of patient illness severity^19^. A transdiagnostic sample would be required to further explore these associations.

Only the discovery sample was used to investigate the diagnostic-effects for the brain analyses. There remained a significant linear association between GMV and symptom severity group after removing ROP and controlling for CHR and ROD, albeit to a smaller degree. Differences in GMV were observed in the gyrus, cingulate, hippocampus, and cerebellum (Table S9). There was a significant correlation between the original discovery statistical t-map and the controlled statistical t-map, with a slightly stronger voxel-wise correlation (r = .24) than parcel-wise correlation (r = .14, *p* < .001), likely due to smaller clusters in the controlled sample. Reduced sample size and increased covariates may have affected the power and findings of the control analyses. There was also a significant large correlation between the original discovery statistical t-map and the symptom dimension controlled statistical t-map (r = .69, *p <* .001), suggesting robustness against symptom type.

### Figure S15: Comparing clinical differences across symptom severity groups before and after controlling for symptom dimension in the discovery sample

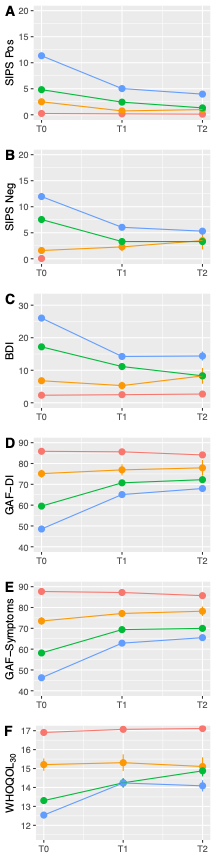

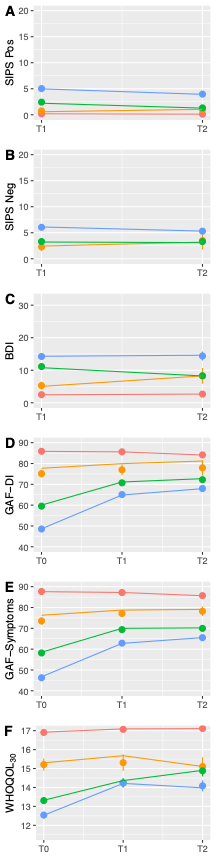

**Uncontrolled	Controlled**

None
Mild
Moderate
Severe

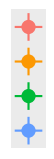

### Figure S16: A) Differences in premorbid functioning from 0-18 years across symptom severity groups and Table S23: B) PAS linear mixed model analyses in the discovery group after controlling for symptom dimension

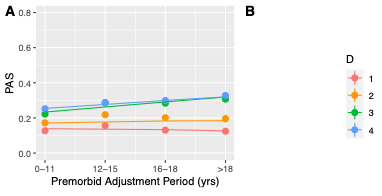

No symptoms
Mild
Moderate
Severe

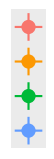

**B**

| **term** | **sumsq** | **SSQ** | **df** | **DenDF** | **F** | **p** | **sig** |
| --- | --- | --- | --- | --- | --- | --- | --- |
| D | 0.243 | 0.081 | 3 | 648.66 | 7.38 | < 0.001 | *** |
| time | 0.337 | 0.337 | 1 | 1,884.70 | 30.79 | < 0.001 | *** |
| BDI | 0.0081 | 0.008 | 1 | 651.50 | 0.74 | 0.39 |  |
| SIPS Positive | 0.025 | 0.025 | 1 | 649.52 | 2.30 | 0.13 |  |
| SIPS Negative | 1.358 | 1.356 | 1 | 650.09 | 123.84 | < 0.001 | *** |
| D:time | 0.547 | 0.182 | 3 | 1,886.41 | 16.62 | < 0.001 | *** |

Notes: D = severity group; meansq = mean square error; DenDF = denominator degrees of freedom; F = F statistic; Rsq = R-squared. Degrees of freedom approximated using Satterthwaite method

**Table S18: Kruskal-Wallis tests for baseline clinical variables across symptom severity groups in the replication sample**

| Variable | No symptoms | Mild | Moderate | Severe | F/Chi2(SD) | Sig | Eta2/Phi |
| --- | --- | --- | --- | --- | --- | --- | --- |
| N | 151 | 24 | 126 | 264 |  |  |  |
| Clinical High Risk, N(%) | 0(0.0) | 0(0.0) | 65(51.6) | 67(25.4) | 110.01(8) | 1.09e-23 | 0.44 |
| Recent Onset Depression, N(%) | 0(0.0) | 15(62.5) | 58(46.0) | 51(19.3) | 109.23(8) | 1.61e-23 | 0.44 |
| Recent Onset Psychosis, N(%) | 0(0.0) | 0(0.0) | 0(0.0) | 145(54.9) | 222.40(8) | 6.09e-48 | 0.63 |
| Control Participants, N(%) | 151(100.0) | 9(37.5) | 3(2.4) | 1(0.4) | 518.64(8) | 4.35e-112 | 0.96 |
| Age, yrs(SD) | 25.0(5.0) | 24.7(5.7) | 24.8(5.9) | 24.7(6.0) | 1.67(3,561) | -- | -- |
| Gender, male(%) | 64(42.4) | 8(33.3) | 65(51.6) | 128(48.5) | 4.99(12) | -- | -- |
| Education, yrs(SD)) | 15.9(3.1) | 14.9(3.0) | 13.8(2.7) | 14.1(7.5) | 55.07(3,555) | 6.64e-12 | 0.02 |
| Marital status, single(%) | 68(45.0) | 17(73.9) | 85(67.5) | 198(75.6) | 49.54(16) | 1.31e-07 | 0.30 |
| Employment status, unemployed(%) | 3(2.0) | 1(4.2) | 17(13.5) | 52(19.7) | 28.49(8) | 2.87e-06 | 0.22 |
| Family psychosis risk, yes(%) | 3(2.0) | 1(4.2) | 23(18.4) | 39(14.9) | 75.31(36) | 3.33e-07 | 0.37 |
| Birth complications, yes(%) | 14(9.3) | 5(20.8) | 27(21.4) | 57(21.9) | 11.41(8) | 9.69e-03 | 0.14 |
| Head Trauma, yes(%) | 10(6.6) | 4(16.7) | 18(14.3) | 53(20.3) | 14.08(8) | 2.80e-03 | 0.16 |
| SIPS Positive, mean(SD) | 0.1(0.4) | 1.5(1.7) | 5.3(5.2) | 11.9(7.4) | 305.76(3,552) | 5.64e-66 | 0.45 |
| SIPS Negative, mean(SD) | 0.0(0.2) | 2.5(2.7) | 9.5(5.3) | 11.8(7.0) | 317.49(3,551) | 1.63e-68 | 0.47 |
| SIPS Disorganised, mean(SD) | 0.0(0.2) | 1.2(1.4) | 2.6(2.3) | 4.1(3.8) | 229.60(3,544) | 1.69e-49 | 0.28 |
| SIPS General, mean(SD) | 0.1(0.5) | 3.2(3.8) | 7.6(3.9) | 8.0(4.5) | 283.51(3,545) | 3.68e-61 | 0.47 |
| SIPS Total, mean(SD) | 0.3(0.9) | 8.5(7.2) | 24.8(11.1) | 35.9(14.9) | 366.32(3,535) | 4.36e-79 | 0.64 |
| SANS Total, mean(SD) | NaN(NaN) | 12.1(13.2) | 23.5(20.7) | 33.4(25.2) | 20.99(2,343) | 2.77e-05 | NaN |
| BDI Total, mean(SD) | 2.6(3.2) | 8.9(5.4) | 19.5(7.5) | 25.8(13.5) | 279.75(3,491) | 2.40e-60 | 0.50 |
| GAF social past month, mean(SD) | 87.9(5.6) | 70.0(13.4) | 51.7(12.8) | 44.4(14.4) | 350.05(3,560) | 1.46e-75 | 0.70 |
| GAF disability past month, mean(SD) | 87.6(5.7) | 72.3(14.2) | 51.8(13.7) | 46.3(13.9) | 343.83(3,560) | 3.23e-74 | 0.68 |
| WHO QoL Total | 15.4(1.5) | 13.4(2.4) | 12.6(1.6) | 12.1(2.1) | 192.86(3,469) | 1.47e-41 | 0.39 |
| WAIS-V Vocabulary | 12.6(2.6) | 10.8(2.7) | 10.9(3.0) | 10.3(4.1) | 53.15(3,537) | 1.71e-11 | 0.07 |
| WAIS-V Matrix Reasoning | 12.0(2.0) | 10.8(2.3) | 10.6(2.7) | 10.5(4.0) | 45.40(3,538) | 7.59e-10 | 0.04 |

1.	Dwyer DB, Buciuman M-O, Ruef A, et al. Clinical, brain, and multilevel clustering in early psychosis and affective stages. *JAMA psychiatry*. 2022;79(7):677-689.

2.	Hastie T, Tibshirani R. Classification by pairwise coupling. *Advances in neural information processing systems*. 1997;10

3.	Rolls ET, Huang CC, Lin CP, Feng J, Joliot M. Automated anatomical labelling atlas 3. *Neuroimage*. Feb 1 2020;206:116189. doi:10.1016/j.neuroimage.2019.116189

4.	Gaser C, Dahnke R, Thompson PM, Kurth F, Luders E, Initiative AsDN. CAT: a computational anatomy toolbox for the analysis of structural MRI data. *Gigascience*. 2024;13:giae049.
